# Estimated Impact of 2022-2023 Influenza Vaccines on Annual Hospital Burden in the United States

**DOI:** 10.1101/2025.03.14.25323919

**Authors:** Kaiming Bi, Shraddha Ramdas Bandekar, Anass Bouchnita, Annalise Cramer, Spencer J. Fox, Rebecca K. Borchering, Matthew Biggerstaff, Lauren Ancel Meyers

## Abstract

During the COVID-19 pandemic early years, infection-prevention measures suppressed transmission of seasonal influenza and other respiratory viruses. The early onset and moderate severity of the US 2022-2023 influenza season may have resulted from reduced use of non-pharmaceutical interventions or lower population immunity after two years of limited influenza virus circulation. We used a mathematical model of influenza virus transmission that incorporates vaccine-derived protection against both infection and severe disease, observed hospitalization burden, to estimate the impact of influenza vaccines on healthcare burden. Despite limited data on vaccine effectiveness against infection, our analyses suggest substantial indirect protection, particularly from young adults to other age groups. This is supported by a significant negative correlation between young adult (aged 18-49 years) vaccination rates and observed hospital burden across US states. Assuming reported levels of past vaccine effectiveness against infection and hospitalization, we estimate that influenza vaccines prevented 67,931 [95% confidence interval (CI): 34,182, 95,842] influenza-related hospitalizations nationwide during the 2022-2023 season, with 61% attributable to reduced susceptibility and onward transmission. Among those aged >=65 years, nearly half of averted hospitalizations resulted from vaccinating younger age groups. These findings highlight the need for better estimates of influenza vaccine effectiveness against infection and the potential benefits of increasing young adult influenza vaccination rates to reduce both direct and indirect disease burden.

**Significance Statement:** Annual influenza epidemics in the United States result in hundreds of thousands of hospitalizations. Quantifying the impact of influenza vaccines in reducing the burden of influenza is crucial, yet many analyses fail to consider the dual benefits of vaccines: directly protecting recipients and indirectly protecting their infectious contacts. Using a mathematical model that accounts for these effects, we estimate that influenza vaccines prevented nearly 68,000 hospitalizations during the 2022-2023 season, with an additional 26,500 hospitalizations potentially avoidable if coverage reached the national target of 70%. Although considerable uncertainty remains about the effectiveness of influenza vaccines in preventing infection, our findings suggest that vaccinating younger adults may offer significant indirect protection against influenza for older adults. Tailoring vaccine campaigns by both age group and US state could further enhance the public health impact of annual vaccination efforts.

## Introduction

Influenza activity in the United States was largely suppressed during the first two years of COVID-19 pandemic non-pharmaceutical interventions and then returned to pre-pandemic levels in late 2022 (1, 2). The early timing and moderate severity of the 2022-2023 influenza season may have been influenced by the circulating influenza A(H3N2) virus subtype (3), reduced protective behaviors like mask-wearing and physical distancing, or lower immunity levels after two years of suppressed transmission (4). Since 2010, seasonal influenza vaccines have been recommended for everyone aged six months and older by the US Centers for Disease Control and Prevention (CDC) and Advisory Committee on Immunization Practices (ACIP), as a means to reduce the risks of infection, severe illness, and transmission to others (5, 6). Influenza vaccination coverage has been slowly increasing, from 41.8% during the 2011-2012 season to 49.3% in the 2022-2023 season (7). In 2020, the Department of Health and Human Services (HHS) set a goal of reaching 70% coverage by 2030, as part of the national Healthy People initiative (8).

Estimating the impact of influenza vaccines on disease burden can be difficult, given that their effectiveness varies annually and across different population groups and that vaccine-acquired reduction in susceptibility can indirectly reduce risks to many others, depending on health-related behavior and social contact patterns (9, 10). Prior studies estimated that annual influenza vaccination averted 7,700 to 40,400 hospitalizations per season from 2005 to 2011 (11), 79,000 hospitalizations during the 2012-13 season (12), 90,000 hospitalizations during 2013-14 season (13), and 58,000 hospitalizations during 2018-19 season (14). One study estimated that increasing influenza vaccine coverage by 5% would have prevented 11,000 hospitalizations during the 2017-18 season (15). For the 2022-2023 season, CDC estimated that influenza vaccination prevented 65,000 hospitalizations in the US. Like many such estimates, these values consider only vaccine effectiveness (VE) in reducing disease severity (e.g., preventing medically-attended illness), and not VE in reducing susceptibility (16).

Here, we estimate the burden of influenza hospitalizations averted during the 2022-2023 season using an age-stratified compartmental model of influenza virus transmission that incorporates humidity-driven seasonality and explicitly tracks the impacts of vaccine- and infection-acquired immunity on both infection susceptibility and disease severity. Our analysis focuses on the first post-COVID pandemic period, which may differ from prior seasons in the extent of infection-acquired immunity, adoption of precautionary behaviors such as face mask-wearing, and the effectiveness and uptake of influenza vaccines (17, 18).

## Results

Between October 1, 2022 and April 29, 2023, 51% of the US population received an influenza vaccine (19) and 209,972 laboratory-confirmed influenza hospital admissions (2) were reported in the United States (not all influenza hospital admissions were reported). The effectiveness of influenza vaccines at preventing infection is much more difficult to measure and is reported far less frequently than their effectiveness at preventing medically attended illness. To estimate the vaccine-averted burden, we assume VE against influenza hospitalization as reported for the 2022–2023 influenza season (20) and VE against infection as reported from a systematic review of VE estimates from medically attended illnesses from earlier A(H3N2)-dominated seasons in the Northern Hemisphere (21) (**Table 1**). Based on these assumptions, we estimate that without vaccines, there would have been an additional 67,931 [95% CI: 34,182, 95,842] influenza hospitalizations during that time period (**Figure 1A**). Approximately 40% of hospitalizations averted (26,506 [95% CI: 9,098, 48,783]) are attributable to the direct protection vaccines provide against severe outcomes; the remainder result from vaccine-related reductions in susceptibility and infectiousness, which directly protect recipients and indirectly protect their social contacts (**Supplementary Table S.A.1)**. Across age groups, adults aged 65 and older had the largest estimated *proportional* reduction in hospitalizations of 32.4% [95%CI: 27.5%, 41.1%], and the largest estimated *per capita* reduction of 80.4 [95%CI: 33.5, 125.3] averted per 100,000 people (**Figure 1B** and **Supplementary Table S.A.2**). Children aged 0 to 17 had the second largest estimated reduction in hospitalizations, with a proportional reduction of 26.1% [95%CI: 19.9%, 39.3%] and an absolute reduction of 15.9 [95%CI: 8.7, 21.3] hospitalizations averted per 100,000 people). While young adults aged 18 to 49 had both the lowest vaccination rate (35.8%) and the smallest estimated reduction in burden (14.8% [95%CI: 9.3%, 25.8%] and 5.0 [95%CI: 3.1, 6.9] per 100,000 people), vaccination of this group is estimated to have indirectly prevented 17.2 [95%CI: 9.6, 20.3] hospitalizations per 100,000 people aged 65 and older **(Figure 1C and Supplementary Table S.C.1)**.

**Table 1:**
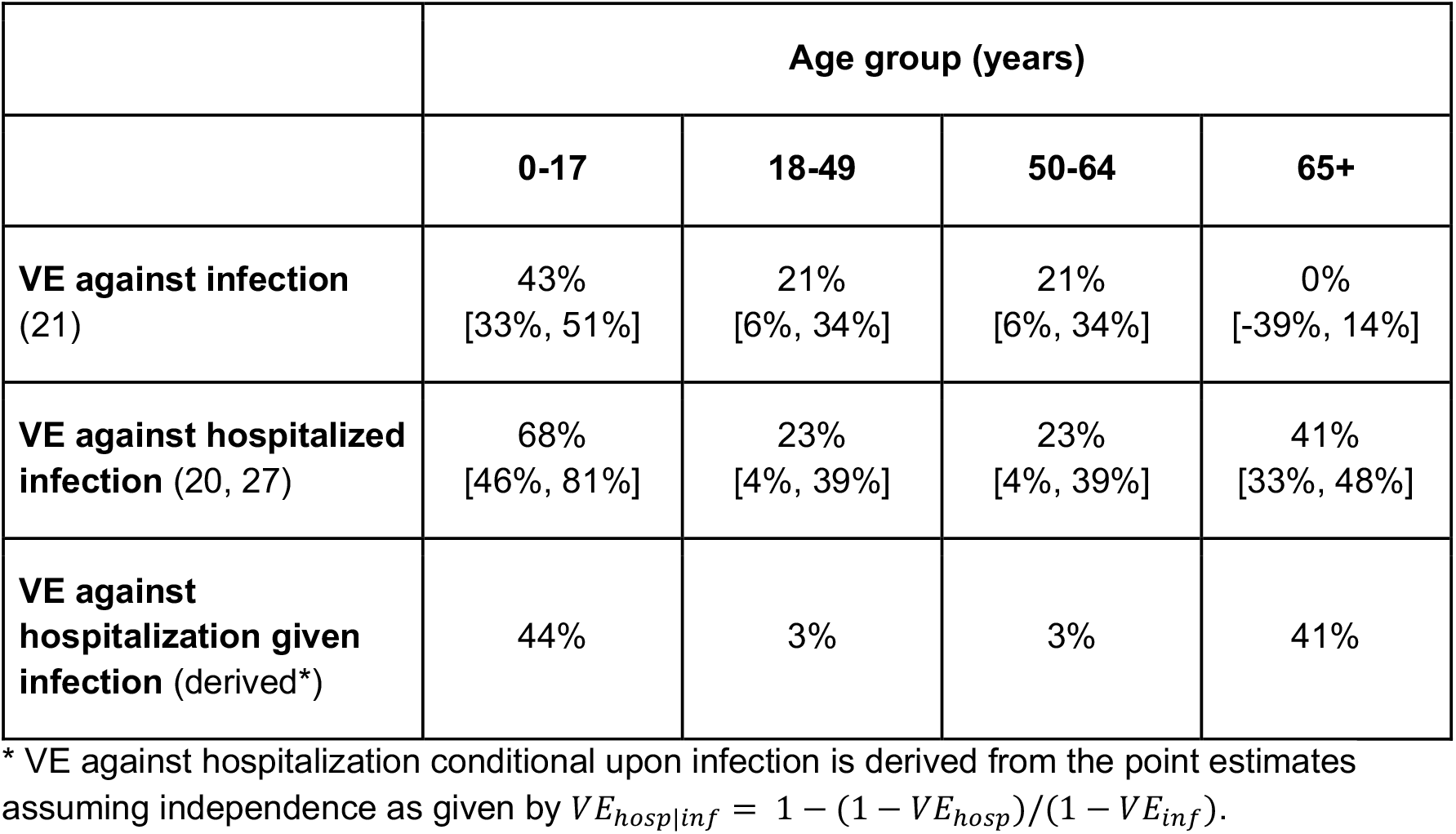
Assumed age-specific vaccine effectiveness (VE) against infection, influenza hospitalized-infection, and hospitalization conditional upon infection during the 2022-2023 influenza season. Values are point estimates and 95% confidence intervals. The primary analysis assumes the point estimates; a supplementary sensitivity analysis considers uncertainty (**Supplementary Information Table S.A.4)**.

**Figure 1:**
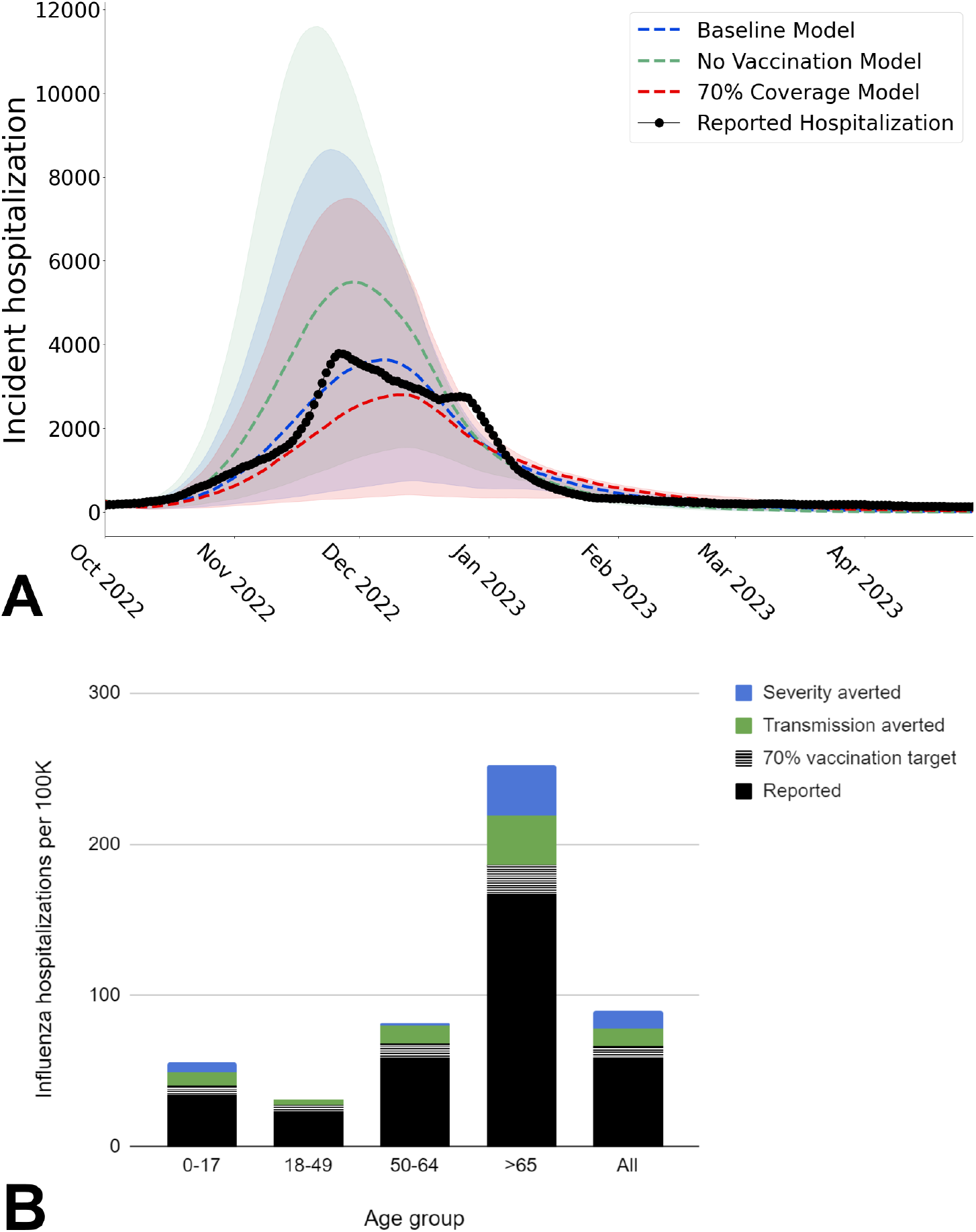
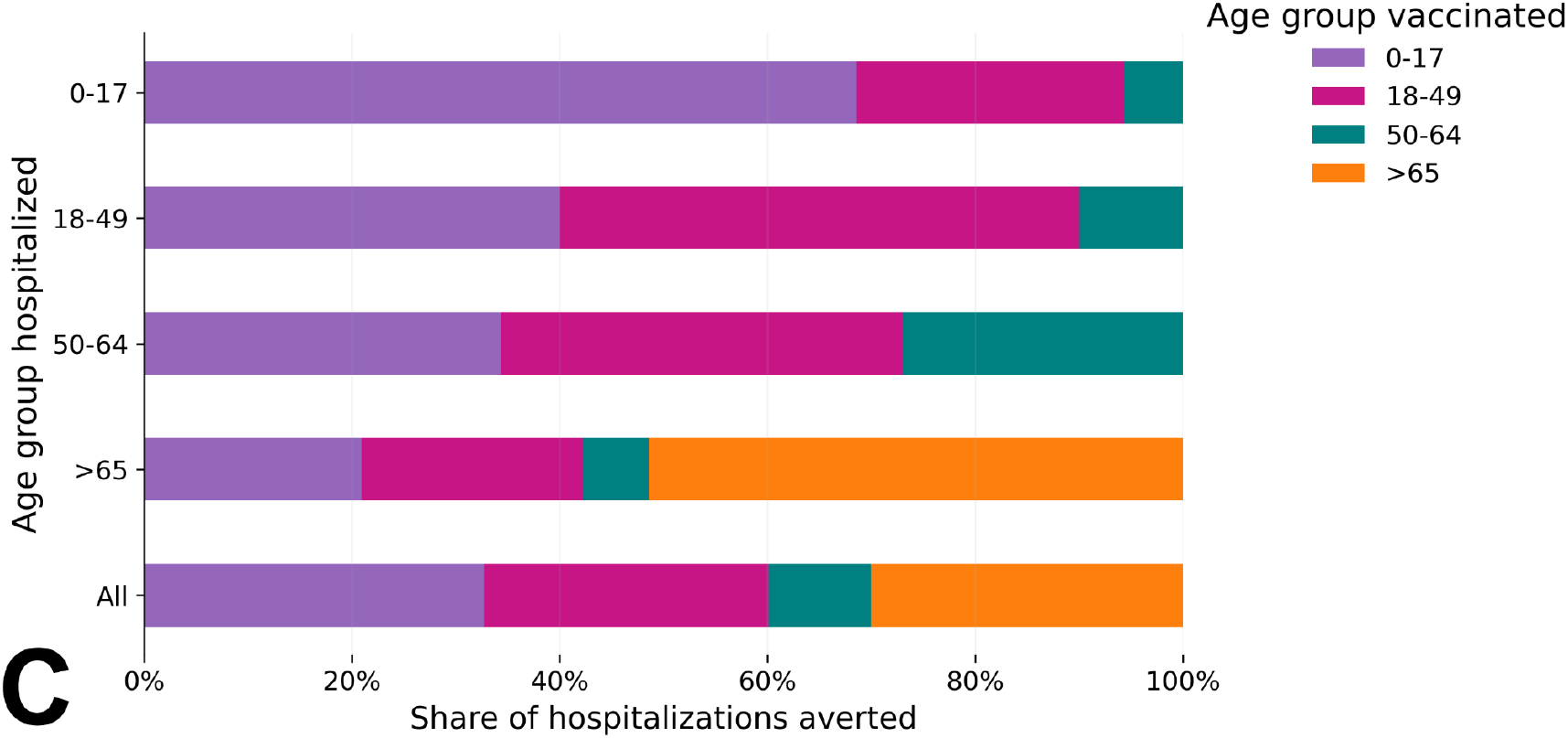
Estimated influenza hospitalizations averted by vaccines from October 1, 2022 to April 29, 2023 in the United States. (A) The estimated seven-day average number of influenza-related hospital admissions in a *baseline scenario* which assumes the reported vaccine coverage (blue), a *no vaccination* counterfactual scenario (green), and an *elevated vaccination* counterfactual scenario which assumes 70% coverage in all age groups (red). Dashed lines and shaded ribbons represent the median and 95% estimation intervals across 1,000 stochastic simulations, respectively. Black dots indicate the reported seven-day average influenza hospital admissions (2). (B) Estimated influenza hospitalizations averted attributable to vaccine-acquired reduction in hospitalization given infection (blue) and vaccine-acquired reduction in susceptibility to infection (green) for each age group during the 2022-2023 influenza season. The black bars indicate the reported hospitalizations and the striped black and white bars indicate the estimated fraction of those hospitalizations that would have been averted with 70% coverage. (C) For each age group, the proportion of influenza hospitalizations averted due to vaccination in different age groups. For example, the top bar indicates that 68.6%, 25.8%, and 5.7% of hospitalizations prevented among children age 0 to 17 stemmed from vaccination of 0 to 17 year olds, 19 to 49 year olds, and 50 to 64 year olds, respectively (**Supplementary Table S.A.2**). The proportions are based on median values of the share of hospitalizations averted across 1,000 stochastic simulations.

To reach the Health People 2030 (22) target of 70% influenza vaccine coverage in all age groups, 77,568,614 more people would have had to receive vaccines in the United States, 69% of whom are young adults aged 18 to 49. Under that scenario, we estimate that there would have been 26,508 [95% CI: 17,228, 33,353] fewer hospitalizations and that roughly 71% of that reduction would stem from the increased influenza vaccine coverage in adults aged 18 to 49 (**Figure 1A and Supplementary Table S.A.3**).

Across US states, reported influenza hospitalizations during the 2022-2023 season ranged from 18.16 to 166.30 per 100,000 people and vaccine uptake ranged from 37.64% to 65.42% overall and 21.12% to 46.64% among young adults ages 18 to 49 (**Figure 2A**). We find that influenza hospitalization burden is significantly correlated with young adult vaccination rates (*p* < 0.0107), and not significantly correlated with the overall vaccination rates (p < 0.0553) (Table S.A.7), and we estimate that a one percentage point decrease in coverage among young adults corresponds to an additional 1.6 influenza hospitalizations per 100,000 (**Figure 2**). Several states deviate considerably from this trend, perhaps due to environmental, health, social, testing, reporting rate or other factors not considered in our analysis.

**Figure 2:**
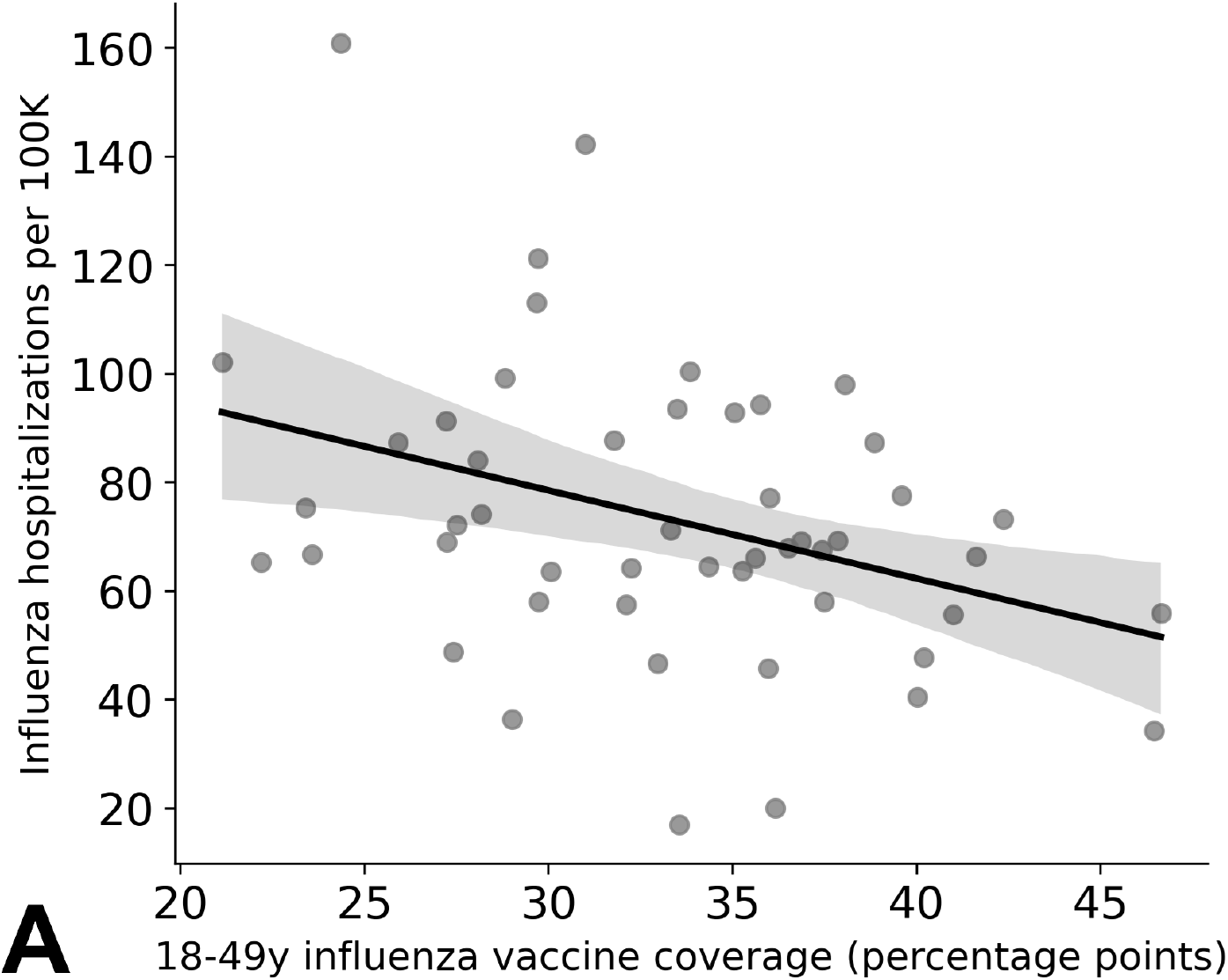
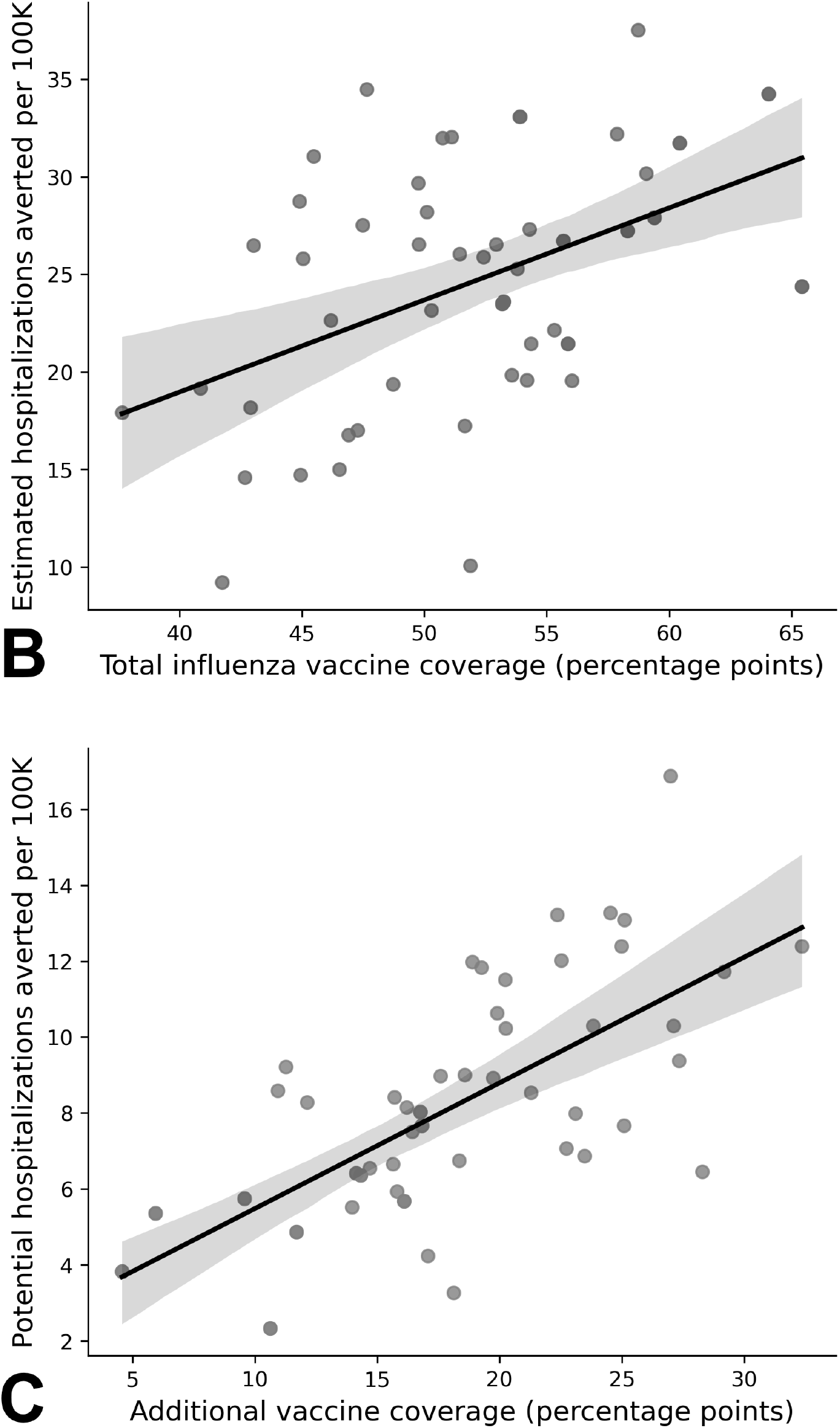
Estimated influenza hospitalizations averted by vaccines from October 1, 2022 to April 29, 2023 across US states. (A) Reported influenza-related hospitalizations per 100,000 are significantly correlated with vaccination rates in the 18-49 year age group across US states. (B) State-level estimates for influenza hospitalizations averted by vaccines significantly correlate with overall vaccine coverage. (C) State-level estimates for additional influenza hospitalizations that would have been averted with 70% coverage significantly correlate with percentage point change in vaccine coverage needed to achieve 70%. Dots correspond to individual states; lines and shading indicate fitted regression lines and their 95% confidence bands, respectively.

We estimate that influenza vaccines averted between 9.2 [95%CI: 5.9 - 14.2] and 37.5 [95%CI: 25.8 - 43.6] influenza hospitalizations per 100,000 across US states (**Figure 2B**). Under the hypothetical 70% coverage scenario, we would expect state-level influenza hospitalizations to have been between 2.08 [95%CI: 1.6-2.3] and 16.9 [95%CI: 12.5 - 25.6] per 100,000 lower, with less pronounced geographic trends (**Figure 2C**). Interestingly, the estimated number of hospitalizations actually averted by 2022-2023 vaccines and the number potentially averted had states reached 70% coverage are not significantly correlated (**Supplementary Figure S.A.1**).

## Discussion

Although influenza vaccination is the most effective way to prevent seasonal influenza (7), vaccination rates in the United States remain far below the national target of 70% in most age groups and vary considerably across states. Counterfactual simulations allow us to estimate the public health impacts of past vaccination campaigns and evaluate the potential direct and indirect impacts of increasing uptake of influenza vaccine in particular subpopulations. Our analyses suggest that, without influenza vaccines, there would have been roughly 25% more influenza hospitalizations during the 2022-2023 influenza season. Alternatively, if vaccination rates had reached the national target, we would expect the influenza hospitalization burden to have been 13% lower than reported.

Our findings are highly sensitive to assumptions about vaccine effectiveness and are limited by the lack of direct data on VE against infection in the 2022–2023 season. Nonetheless, our estimate of 67,931 [95% CI: 34,182, 95,842] influenza hospitalizations averted by vaccines aligns with prior seasonal estimates, ranging from 7,700 in the 2009–2010 season (11) to 90,000 in the 2013-2014 influenza season (13). Several factors may have contributed to the 2022-2023 estimates being at the higher end of this range. First, the vaccine coverage was 2% to 8% higher than in all seasons prior to 2019-2020 (23, 24). Second, the influenza A(H3N2) virus subtype––which dominated the 2022-2023 season––typically causes a higher hospitalization burden than A(H1N1) (25). Third, the 2022-2023 season unfolded following years of COVID-19-related suppression of influenza virus transmission (26), as the use of non-pharmaceutical prevention measures was declining.

Although our estimate is consistent with CDC’s estimate of 65,000 influenza hospitalizations averted in 2022-2023 season from influenza vaccination (16), we note three key methodological differences. First, unlike most prior estimates, we consider the effectiveness of vaccines at reducing susceptibility to infection in addition to their effectiveness at reducing the risks of severe outcomes once infected. Second, we assumed vaccine effectiveness (VE) estimates reported by the Virtual SARS-CoV-2, Influenza, and Other respiratory viruses (VISION) network for adults (27), whereas CDC assumed higher VE pooled from multiple inpatient and outpatient samples (28). Finally, the CDC estimate adjusts for underreporting of laboratory-confirmed influenza hospital admissions.

National influenza vaccination campaigns have prioritized older adults, who are at increased risk for severe disease, and children, who are at increased risk for infection (30). Our results corroborate the substantial public health benefits of this strategy and suggest that the Advisory Committee on Immunization Practices (ACIP) 2024–25 influenza vaccine recommendations, which include higher dose or adjuvanted vaccines for older adults (31), might further reduce burden in this age-group.

We also find that the vaccination of young adults, ages 18 to 49, could provide indirect protection again influenza for other age groups, particularly older adults, and that increasing vaccination rates in 18-49 age group could reduce the annual burden of influenza. When we analyzed vaccination and hospitalization rates across all 50 US states, we observed a significant correlation between influenza hospitalizations and vaccine uptake in young adults, but not with total vaccine coverage among all age groups. The population-wide benefits of preventing infection in young adults stems from both their large representation (40% of the US population) and their high interaction rates with other age groups (32). Given that young adults have the lowest vaccination rates compared to other age groups (37.49% in 2022-2023), there is substantial opportunity to boost coverage in this age group.

Our findings rest on several modeling assumptions with considerable uncertainty, including immunity from prior influenza infections, vaccine effectiveness, immune waning rates, and mixing rates between age groups—particularly given lasting behavioral changes from the COVID-19 pandemic that have not been fully measured. Additionally, our model is fitted to hospitalization admission data, which does not account for unobserved or missing reported events, and we did not account for variation in hospital testing practices or hospitalization reporting rates across age groups or states. Our assumptions about vaccine effectiveness against infection are based on a published systematic review of data from multiple influenza seasons across the Northern Hemisphere (21), whereas assumptions regarding vaccine effectiveness against hospitalization are derived from studies of the 2022–2023 season in the United States (27).

Our study uses dynamic, data-driven models to disentangle the dual benefits of vaccines–– preventing infections and reducing disease severity—while identifying key age groups for targeted interventions. With improved estimates of vaccine protection against influenza infection, this approach could better clarify the direct, indirect, and varied benefits of vaccination, guiding more effective public health communication and immunization strategies.

## Materials and Methods

We estimated hospitalizations averted by influenza vaccines during the 2022-2023 influenza season using an age-stratified stochastic compartmental model of influenza virus transmission that includes humidity- and school-related seasonality and tracks changing levels of infection-acquired and vaccine-acquired immunity against both infection and hospitalizations (33). We fit the model to hospitalization data (2) to estimate the transmission rate and set baseline influenza vaccination roll-out to match reported vaccination patterns (19). We used the model to simulate the *baseline* scenario (i.e., 2022-2023 vaccination rates) and five types of counterfactual scenarios: (i) no vaccination, (ii) target vaccination (70% vaccine coverage across all age groups), (iii) vaccination of a single age group only, at the observed 2022-2023 rate, (iv) vaccination of a single age group only, at the target 70% rate, and (v) vaccines protect against infection only (i.e., vaccine effectiveness against severity upon infection is set to zero).

To estimate the influenza hospitalizations averted by a given scenario, we performed pairwise comparisons between simulations of that scenario and simulations of the *baseline* scenario. Specifically, to estimate the influenza hospitalization burden averted as a result of the 2022-2023 vaccination campaign, we compare the *baseline* scenario to the *no vaccination* scenario. To further estimate the impact of vaccines in a single age group, we compare the corresponding single-group vaccination scenario to the *no vaccination* scenario. To project the additional burden averted had 70% population been vaccinated (overall or in a specific age group), we compare the specified target vaccination scenario with the *baseline* scenario. Finally, to disentangle the two effects of vaccines—reducing susceptibility and reducing severity—we compare a scenario where vaccines only protect against infection (with vaccine effectiveness against severity set to zero) to both the *no vaccination* scenario and the *baseline* scenario. The first comparison allows us to estimate the population-level impact of vaccine effectiveness in preventing infection, while the second indicates the impact of vaccine effectiveness against severity.

We conducted these scenario comparisons for the entire US and each of the 50 states. For each comparison, we ran 1,000 pairs of simulations that shared fixed random seeds.

### Data

Assumed influenza vaccination rates and hospital admissions are based on publicly available data from the CDC (19) and US Department of Health & Human Services (2), respectively.

Demographic parameters are based on US Census Bureau reports (34). Absolute humidity time series for each state and the US are from the National Oceanic and Atmospheric Administration (NOAA) (35). The school and work calendar is derived from data available from the US Department of Commerce (36) and the Department of Education (37). We refer the reader to the Supplementary Information for more details.

### Epidemic model

Our age-structured SEIRS (Susceptible-Exposed-Infectious-Recovered-Susceptible) model of influenza transmission includes six age groups (0-4, 5-11, 12-17, 18-49, 50-64, and 65+) and explicitly tracks the presumed immunity resulting from infection by either an influenza A(H1N1) or influenza A(H3N2) virus subtype and vaccination followed by waning (**Figure 3B**). Infected individuals are hospitalized according to age-specific infection hospitalizations rates (38, 39). Population-wide levels of immunity impact both susceptibility to and severity of infections, modulated by age-specific vaccine uptake (19) and effectiveness (21, 28, 40) parameters. We refer the reader to **Supplementary Information B** for more details.

**Figure 3:**
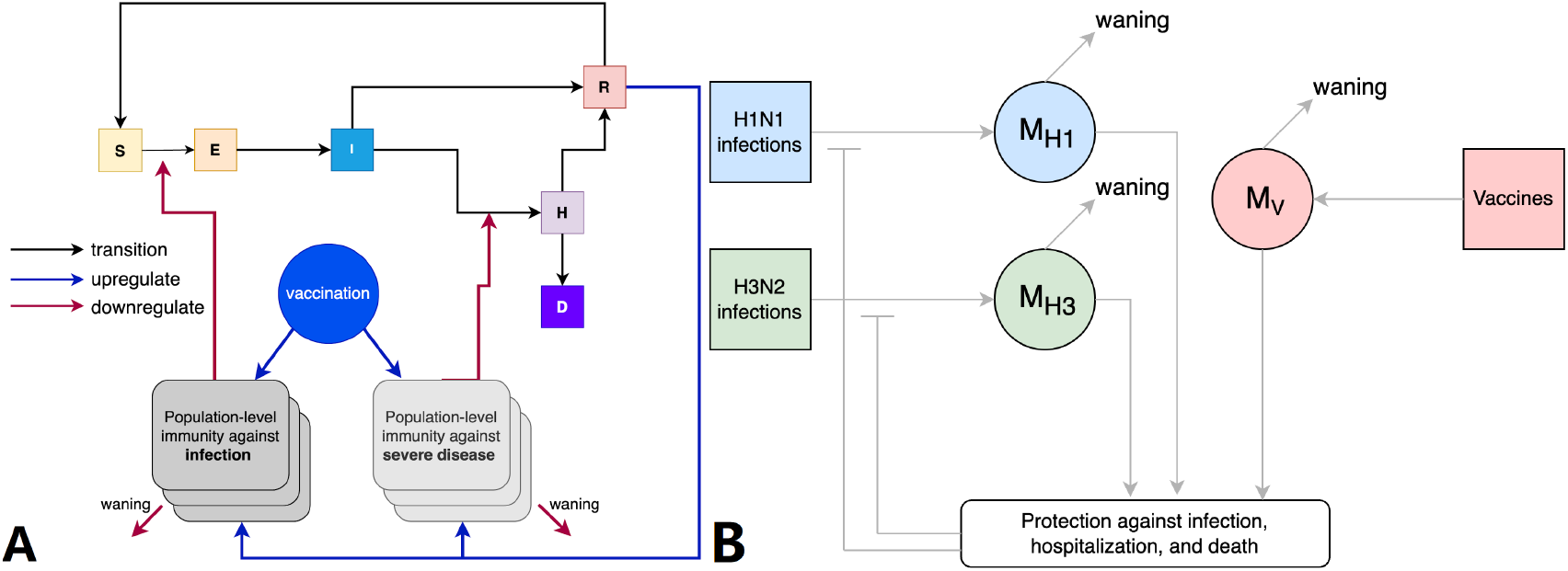
Schematic representation of influenza virus transmission model that tracks population-level immunity against two influenza subtypes derived from natural infection and vaccination for six age groups. **(A)** Susceptible individuals (*S*) move to the exposed state (*E*) upon infection. Exposed individuals transition to the infectious compartment (*I*); a fraction of individuals move directly from (*I*) to the recovered compartment (*R*), while the remaining transition to the hospitalized compartment (*H*). Hospitalized cases either recover (*R*) or die (*D*). Recovered individuals eventually become partially susceptible again and move into the susceptible compartment (*S*). For each type of immune exposure (i.e., infection with a specific subtype or receipt of vaccine), the model uses two state variables to track the resulting population-level average protection against infection and severe disease. These variables increase as individuals recover from infections and receive vaccines and they decrease according to waning (half-life) parameters, specific to each exposure type. Immunity state variables modify overall rates of infection and risk of hospitalization/death with effectiveness that can vary depending on currently circulating virus subtypes and the age group of the exposed individual. **(B)** Infection upregulates population immunity depending on the relative frequency of each subtype among active infections. These are given by *M*_*H*1_ and *M*_*H*3_. The administration of vaccine increases vaccine-derived immunity, represented as *M*_V_. We model a separate set of infection and immunity variables for each of the following age groups: 0-4, 5-11, 12-18, 19-49, 50-64, and 65+.) move to the exposed state upon infection. Exposed individuals transition to the infectious compartment ; a fraction of individuals move directly from to the recovered compartment, while the remaining transition to the hospitalized compartment. Hospitalized cases either recover or die. Recovered individuals eventually become partially susceptible again and move into the susceptible compartment. For each type of immune exposure (i.e., infection with a specific subtype or receipt of vaccine), the model uses two state variables to track the resulting population-level average protection against infection and severe disease. These variables increase as individuals recover from infections and receive vaccines and they decrease according to waning (half-life) parameters, specific to each exposure type. Immunity state variables modify overall rates of infection and risk of hospitalization/death with effectiveness that can vary depending on currently circulating virus subtypes and the age group of the exposed individual. **(B)** Infection upregulates population immunity depending on the relative frequency of each subtype among active infections. These are given by *M*_*H*1_ and *M*_*H*3_. The administration of vaccine increases vaccine-derived immunity, represented as *M*_V_. We model a separate set of infection and immunity variables for each of the following age groups: 0-4, 5-11, 12-18, 19-49, 50-64, and 65+.

Our model tracks both inter-seasonal immunity that decreases rapidly at the onset of a new season and intraseasonal immunity that increases with new infections and vaccinations. Based on the influenza-positive test data from FluView (25), both the 2021-2022 and 2022-2023 influenza seasons were A(H3N2)-dominated. Therefore, we assumed that both the inter-seasonal and intra-seasonal immunity are contributed to by the A(H3N2) subtype. In our model, we consider both the population-level immunity resulting from A(H3N2) infections against infection, and against hospitalization. Similarly, we consider both the population-level immunity derived from vaccines against infection, and against hospitalization. We refer the reader to **Supplementary Information B.3** for more details.

The model incorporates humidity-driven seasonality, based on normalized absolute humidity levels for each state derived from NOAA data (35), as well as calendar-driven seasonality, based on age-specific mixing matrices for specific venues (32) and national school (41) and holiday (36) data. We refer the reader to **Supplementary Information B.4** and **C.3** for more details.

### Model Calibration

The following model parameters are calibrated through least square fitting of the model with the baseline scenario to the seven-day average of influenza hospitalizations between October 1, 2022 and April 29, 2023 (2), at both the state and national level. For the other counterfactual scenarios, the model adopts the calibrated parameters from the baseline scenario. We used the Python curve-fit function (42), which implements the least squares solver through the Scipy (43). The calibrated parameters are provided in **Table S.B.4**.

### Vaccine Effectiveness

The assumed age-specific effectiveness of vaccines against infection (i.e., effectiveness at reducing susceptibility), hospitalized infection, and hospitalization conditional upon infection (i.e., effectiveness at reducing severity) are provided in **Table 1**. For effectiveness against hospitalized infection, we use estimates obtained by the New Vaccine Surveillance Network (NVSN) for children under 18 years (28) and by the Virtual SARS-CoV-2, Influenza, and Other respiratory viruses Network (VISION) Network for adults (27), (28). For effectiveness against infection, we relied on a published meta-analysis which estimated effectiveness against symptomatic infection stratified by season, influenza subtype, and age group from 56 studies between 2006 and 2015 (21). We recomputed pooled age-stratified estimates based on nine studies from the Northern Hemisphere in which influenza A(H3N2) was the dominant seasonal subtype. Finally, we calculate vaccine effectiveness against hospitalization conditional upon infection from the point estimates assuming independence (44) as given below **Table 1**.

### Counterfactual simulation experiments

For each scenario-based estimate, we used the corresponding model to simulate 1,000 epidemics, incorporating both parameter uncertainty and process stochasticity. Whenever comparing between scenarios, we ran 1,000 pairs of simulations that each shared a random seed. For each run, the transmission rate and the seasonality coefficient are sampled from normal distributions with the corresponding calibrated mean and standard deviation.

Stochasticity in disease progression was incorporated using the tau-leaping method (45); transition rates between compartments are sampled from a Poisson distribution with mean equal to the deterministic rates multiplied by the time step. **Supplementary Information B.6** provides additional details. and the seasonality coefficient are sampled from normal distributions with the corresponding calibrated mean and standard deviation. Stochasticity in disease progression was incorporated using the tau-leaping method (45); transition rates between compartments are sampled from a Poisson distribution with mean equal to the deterministic rates multiplied by the time step. **Supplementary Information B.6** provides additional details.

### Sensitivity Analysis

To assess the sensitivity of our results to assumptions about vaccine effectiveness, we analyzed scenarios with VE values set to the upper and lower bounds of the 95% confidence intervals provided in **Table 1. Supplementary Information A.4** provides additional details.

## Data Availability

All data for reproducing the analysis are publicly available at: https://github.com/bikaiming93/Flu_burden_avert

## Acknowledgments

This research was supported by CDC grant U01 IP001136, CSTE grant NU38OT000297, and CDC CFA NU38FT000008.

## Reference

1. Website. Available at: CDC. Influenza Activity in the United States during the 2022–23 season and Composition of the 2023–24 influenza vaccine. Centers for Disease Control and Prevention https://www.cdc.gov/flu/spotlights/2023-2024/22-23-summary-technical-report.htm (2023). [Accessed 2 May 2024].

2. U.S. Department of Health, Human Services, COVID-19 reported patient impact and hospital capacity by state timeseries (RAW). https://healthdata.gov/Hospital/COVID-19-Reported-Patient-Impact-and-Hospital-Capa/g62h-syeh/about_data [Accessed 2 May 2024].

3. J.-E. Park, Y. Ryu, Transmissibility and severity of influenza virus by subtype. Infect. Genet. Evol. 65, 288–292 (2018).

4. CDC, Influenza Activity in the United States during the 2022–2023 Season and Composition of the 2023–2024 Influenza Vaccine. Influenza (Flu) (2024). Available at: https://www.cdc.gov/flu/whats-new/22-23-summary-technical-report.html [Accessed 28 October 2024].

5. CDC, Prevention and Control of Seasonal Influenza with Vaccines: Recommendations of the Advisory Committee on Immunization Practices — United States, 2024–25 Influenza Season. Centers for Disease Control and Prevention (2024). Available at: https://www.cdc.gov/mmwr/volumes/73/rr/rr7305a1.htm [Accessed 22 Jan 2025].

6. A. Briggs, M. Sculpher, K. Claxton, Decision Modelling for Health Economic Evaluation (OUP Oxford, 2006).

7. CDC, Weekly flu vaccination dashboard. Centers for Disease Control and Prevention (2024). Available at: https://www.cdc.gov/fluvaxview/dashboard/index.html [Accessed 2 May 2024].

8. Increase the proportion of people who get the flu vaccine every year — Data - Healthy People 2030. Available at: https://health.gov/healthypeople/objectives-and-data/browse-objectives/vaccination/increase-proportion-people-who-get-flu-vaccine-every-year-iid-09/data [Accessed 2 May 2024].

9. K. E. C. Ainslie, M. Haber, W. A. Orenstein, Challenges in estimating influenza vaccine effectiveness. Expert Rev. Vaccines 18, 615–628 (2019).

10. A. E. Macias, et al., The disease burden of influenza beyond respiratory illness. Vaccine 39 Suppl 1, A6–A14 (2021).

11. D. Kostova, et al., Influenza Illness and Hospitalizations Averted by Influenza Vaccination in the United States, 2005-2011. PLoS One 8, e66312 (2013).

12. Centers for Disease Control and Prevention (CDC), Estimated influenza illnesses and hospitalizations averted by influenza vaccination - United States, 2012-13 influenza season. MMWR Morb. Mortal. Wkly. Rep. 62, 997–1000 (2013).

13. C. Reed, et al., Estimated influenza illnesses and hospitalizations averted by vaccination--United States, 2013-14 influenza season. MMWR Morb. Mortal. Wkly. Rep. 63, 1151–1154 (2014).

14. J. R. Chung, et al., Effects of Influenza Vaccination in the United States During the 2018-2019 Influenza Season. Clin. Infect. Dis. 71, e368–e376 (2020).

15. M. M. Hughes, et al., Projected Population Benefit of Increased Effectiveness and Coverage of Influenza Vaccination on Influenza Burden in the United States. Clin. Infect. Dis. 70, 2496–2502 (2020).

16. CDC, Preliminary Estimated Flu Disease Burden 2022–2023 Flu Season. Centers for Disease Control and Prevention (2023). Available at: https://www.cdc.gov/flu-burden/php/data-vis/2022-2023.html [Accessed 31 May 2024].

17. M. Sobczak, R. Pawliczak, Which Factors Were Related to the Number of COVID-19 Cases in the 2022/2023 Season Compared to the 2021/2022 Season in Europe? J. Clin. Med. Res. 12 (2023).

18. A. Merced-Morales, Influenza Activity and Composition of the 2022–23 Influenza Vaccine — United States, 2021–22 Season. MMWR Morb. Mortal. Wkly. Rep. 71 (2022).

19. Influenza Vaccine Doses Distributed, United States. (2024). Available at: https://www.cdc.gov/flu/fluvaxview/dashboard/vaccination-doses-distributed.html [Accessed 5 March 2024].

20. Website. Available at: CDC, Flu Vaccine Effectiveness (VE) Data for 2022-2023. Flu Vaccines Work (2024). Available at: https://www.cdc.gov/flu-vaccines-work/php/effectiveness-studies/2022-2023.html [Accessed 12 October 2024].

21. E. A. Belongia, et al., Variable influenza vaccine effectiveness by subtype: a systematic review and meta-analysis of test-negative design studies. Lancet Infect. Dis. 16, 942–951 (2016).

22. Increase the proportion of people who get the flu vaccine every year — IID-09. Available at: https://health.gov/healthypeople/objectives-and-data/browse-objectives/vaccination/increase-proportion-people-who-get-flu-vaccine-every-year-iid-09 [Accessed 13 March 2024].

23. Flu vaccination coverage, United States, 2022–23 influenza season. (2023). Available at: https://www.cdc.gov/flu/fluvaxview/coverage-2223estimates.htm [Accessed 6 May 2024].

24. Website. Available at: Preliminary Flu Vaccine Effectiveness (VE) Data for 2023-2024. (2024). Available at: https://www.cdc.gov/flu/vaccines-work/2023-2024.html [Accessed 6 May 2024].

25. National, regional, and state level outpatient illness and viral surveillance. Available at: https://gis.cdc.gov/grasp/fluview/fluportaldashboard.html [Accessed 1 April 2024].

26. RSV, COVID-19, and flu outlook for 2023-2024. Johns Hopkins Bloomberg School of Public Health (2023). Available at: https://publichealth.jhu.edu/2023/rsv-covid-19-and-flu-outlook-for-2023-2024 [Accessed 6 May 2024].

27. M. W. Tenforde, et al., Influenza Vaccine Effectiveness Against Influenza A-Associated Emergency Department, Urgent Care, and Hospitalization Encounters Among US Adults, 2022-2023. J. Infect. Dis. 230, 141–151 (2024).

28. CDC, Flu vaccine provided substantial protection this season. Centers for Disease Control and Prevention (2023). Available at: https://www.cdc.gov/flu/spotlights/2022-2023/flu-vaccine-protection.htm [Accessed 4 October 2023].

29. CDC, Flu Vaccine Effectiveness (VE) Data for 2022-2023. Flu Vaccines Work (2024). Available at: https://www.cdc.gov/flu-vaccines-work/php/effectiveness-studies/2022-2023.html [Accessed 12 October 2024].

30. Viboud, Cécile, et al. Risk factors of influenza transmission in households. British Journal of General Practice 54.506. 684–689 (2004).

31. L. A. Grohskopf, Prevention and Control of Seasonal Influenza with Vaccines: Recommendations of the Advisory Committee on Immunization Practices — United States, 2024–25 Influenza Season. MMWR Recomm. Rep. 73 (2024).

32. K. Prem, A. R. Cook, M. Jit, Projecting social contact matrices in 152 countries using contact surveys and demographic data. PLoS Comput. Biol. 13, e1005697 (2017).

33. Scenario Projections for SARS-CoV-2, Influenza, and RSV Burden in the US (2023-2024). Available at: https://www.researchsquare.com/article/rs-3467930/latest [Accessed 5 March 2024].

34. United States Census Bureau > Communications Directorate - Center for New Media, QuickFacts: United States.

35. R. D. Cotton, National Oceanic and Atmospheric Administration and the environment. Arch. Environ. Health 22, 404–405 (1971).

36. Holidays. U.S. Department of Commerce. Available at: https://www.commerce.gov/hr/employees/leave/holidays [Accessed 1 April 2024].

37. Home. Available at: https://www.ed.gov/ [Accessed 1 April 2024].

38. National center for health statistics mortality surveillance system. Available at: https://gis.cdc.gov/grasp/fluview/mortality.html [Accessed 12 October 2023].

39. I. Giacchetta, C. Primieri, R. Cavalieri, A. Domnich, C. de Waure, The burden of seasonal influenza in Italy: A systematic review of influenza-related complications, hospitalizations, and mortality. Influenza Other Respi. Viruses 16, 351–365 (2022).

40. M. T. Osterholm, N. S. Kelley, A. Sommer, E. A. Belongia, Efficacy and effectiveness of influenza vaccines: a systematic review and meta-analysis. Lancet Infect. Dis. 12, 36–44 (2012).

41. Home. Available at: https://www.ed.gov/ [Accessed 2 April 2024].

42. curve_fit — SciPy v1.14.0 Manual. Available at: https://docs.scipy.org/doc/scipy/reference/generated/scipy.optimize.curve_fit.html [Accessed 2 July 2024].

43. least_squares — SciPy v1.14.0 Manual. Available at: https://docs.scipy.org/doc/scipy/reference/generated/scipy.optimize.least_squares.html#scipy.optimize.least_squares [Accessed 2 July 2024].

44. M. Elizabeth Halloran, I. M. Longini Jr, C. J. Struchiner, Design and Analysis of Vaccine Studies (Springer Science & Business Media, 2009).

45. S. J. Fox, et al., Real-time pandemic surveillance using hospital admissions and mobility data. Proc. Natl. Acad. Sci. U. S. A. 119 (2022).

