## Supplementary Information for "Estimated Impact of 2022-2023 Influenza Vaccines on Annual Hospital Burden in the United States"

### **Supplement A –– Additional Results**

#### **A.1 Hospitalizations prevented by vaccine-acquired reduction in susceptibility versus vaccine-acquired reduction in severity**

Vaccines can prevent hospitalizations by reducing susceptibility to infection (P_SUS_) or risk of severe disease upon infection in vaccinated individuals (P_SEV_), as well as by reducing risks of transmission to close contacts of vaccinated individuals (P_CON_). To separate the impact of P_SEV_ (*severity* protection) from the combined impact of P_SUS_ and P_CON_ (*infection* protection) in reducing influenza hospitalization during the 2022-2023 season, we ran counterfactual simulations in which vaccines are assumed to protect against infection, with efficacy estimated for 2022-2023, but not against severity (**Table S.A.1**). We compared the projected burdens to those of the *no vaccination* model to assess the impact of *infection* protection, and to those of the *baseline* model to assess the impact of *severity* protection. We estimate that an expected 8.7 [95%CI: 1.9 - 13.7] influenza hospitalizations per 100K in the US were prevented by vaccine-acquired reduction in severity, which translates to roughly 40% of the total hospitalizations averted by vaccines, and that 13.6 [95%CI: 8.6 - 16.5] influenza hospitalizations per 100K in the US were prevented by vaccine-acquired reduction in susceptibility and infectiousness. We also partitioned these two effects of vaccines by age group (**Table S.A.1**).

**Table S.A.1: Estimated influenza hospitalizations averted by 2022-2023 vaccines in each age group, stratified by the two different effects of vaccines (protection against infection and protection against severity if infected).** Percent averted and averted per 100K are the estimated reductions in hospitalizations relative to the number estimated under a *no vaccination* scenario. Values are medians and 95% confidence intervals based on 1,000 pairs of stochastic simulations.

|  | **Percent averted** | | | **Averted per 100K** | | |
| --- | --- | --- | --- | --- | --- | --- |
| **Age group** | **Reduced infection** | **Reduced**  **severity** | **Total** | **Reduced infection** | **Reduced**  **severity** | **Total** |
| **0-17y** | 18%  [13% - 29%] | 8%  [7% - 10%] | 26%  [20% - 39%] | 11.3  [7.3 – 13.1] | 4.6  [1.4– 8.4] | 15.9  [8.7- 21.3] |
| **18-49y** | 14%  [ 10% - 23%] | 1%  [-1% - 3%] | 15%  [9% - 26%] | 4.7  [3.5 – 5.6] | 0.3  [-0.4 – 1.3] | 5.0  [3.1 - 6.9] |
| **50-64y** | 17%  [12% - 28%] | 1%  [0% - 3%] | 18%  [12% - 31%] | 16.1  [10.4 – 18.3] | 1.2  [0.3 - 3.1] | 17.3  [10.7 - 21.4] |
| **> 65y** | 16%  [16% - 15%] | 16%  [12% - 26%] | 32%  [28% - 41%] | 39.8  [20.3 – 49.5] | 40.6  [12.2 – 75.8] | 80.4  [33.5 - 125.3] |
| **All** | 17%  [13% - 25%] | 9%  [8% - 11%] | 25%  [21% - 35%] | 13.6  [8.6 – 16.5] | 8.7  [1.9– 13.7] | 22.3  [11.6 – 31.4] |

#### **A.2 Hospitalizations prevented by age-specific vaccination**

To estimate the impact of vaccinating a single age group on 2022-2023 influenza hospitalization rates in other age groups, we compared the *no vaccination* scenario to scenarios in which only one age group is vaccinated according to the 2022-2023 vaccination rate and all other age groups remain unvaccinated (**Table S.A.2**). To validate these estimates, we also considered a set of *leave-one-unvaccinated* scenarios, in which a single age group is not vaccinated and all the others are vaccinated at 2022-2023 rates. When we estimate burden averted through pairwise comparisons between these scenarios and the *baseline* scenario, we obtained almost identical estimates to those given in **Table S.A.2** (not shown).

**Table S.A.2: Estimated influenza hospitalizations averted by 2022-2023 vaccines in each age group, stratified by age group vaccinated.** Percent averted and averted per 100K are the estimated reductions in hospitalizations relative to the number estimated under a *no vaccination* scenario. Values are medians and 95% confidence intervals based on 1,000 pairs of stochastic simulations.

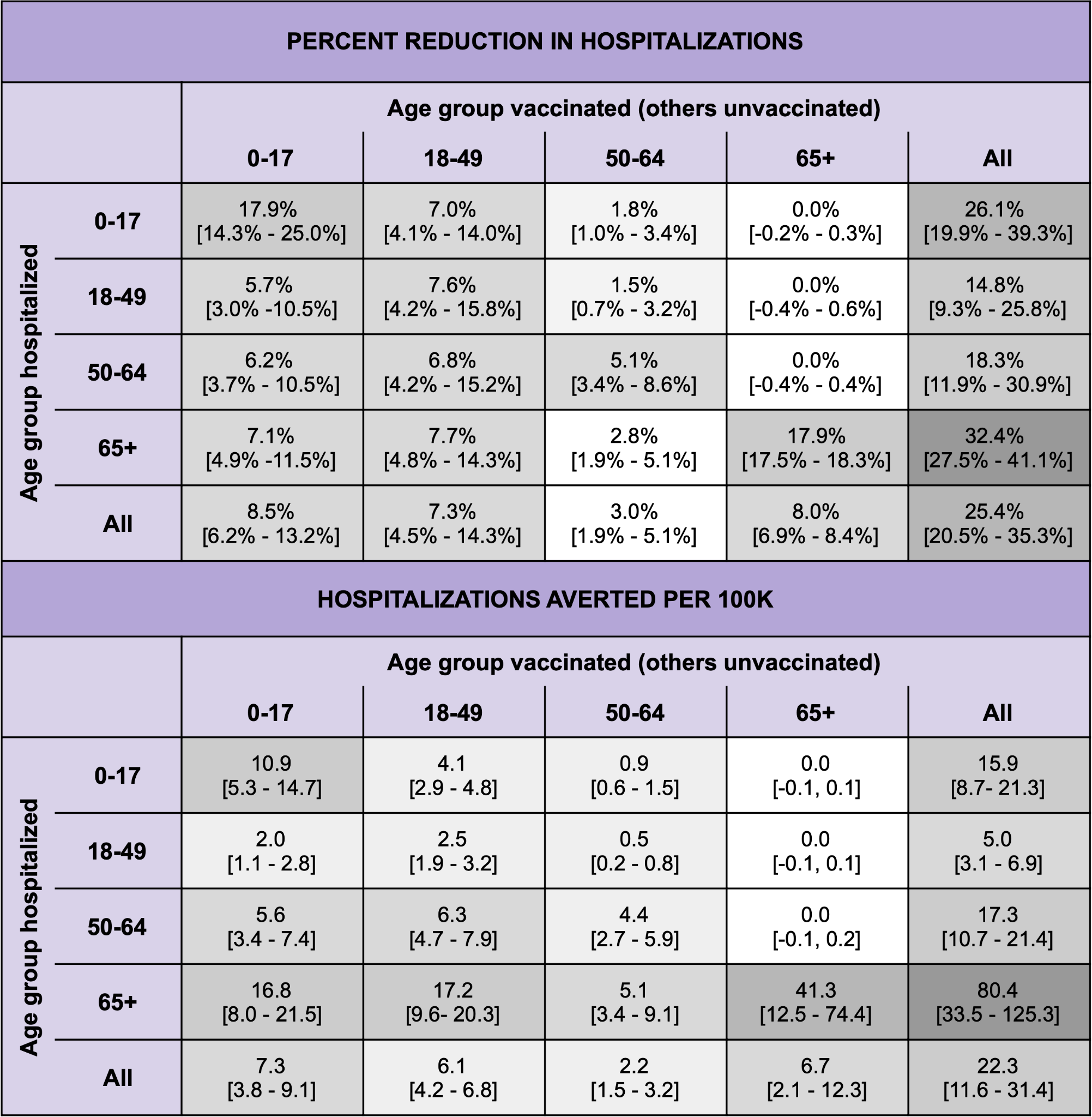

#### **A.3. Hospitalizations potentially averted with 70% vaccination coverage**

To estimate the potential reduction in hospitalizations if 70% of the US population had been vaccinated in each specific age group, we compared the counterfactual scenario in which 70% of all age groups are vaccinated to the *baseline* scenario (**Table S.A.3**).

**Table S.A.3: Estimated *additional* influenza hospitalizations averted if 70% of the US population had received 2022-2023 influenza vaccines.** Percent averted and averted per 100K are the estimated reductions in hospitalizations relative to the *baseline* 2022-2023 vaccination scenario. Values are medians and 95% confidence intervals based on 1,000 pairs of stochastic simulations.

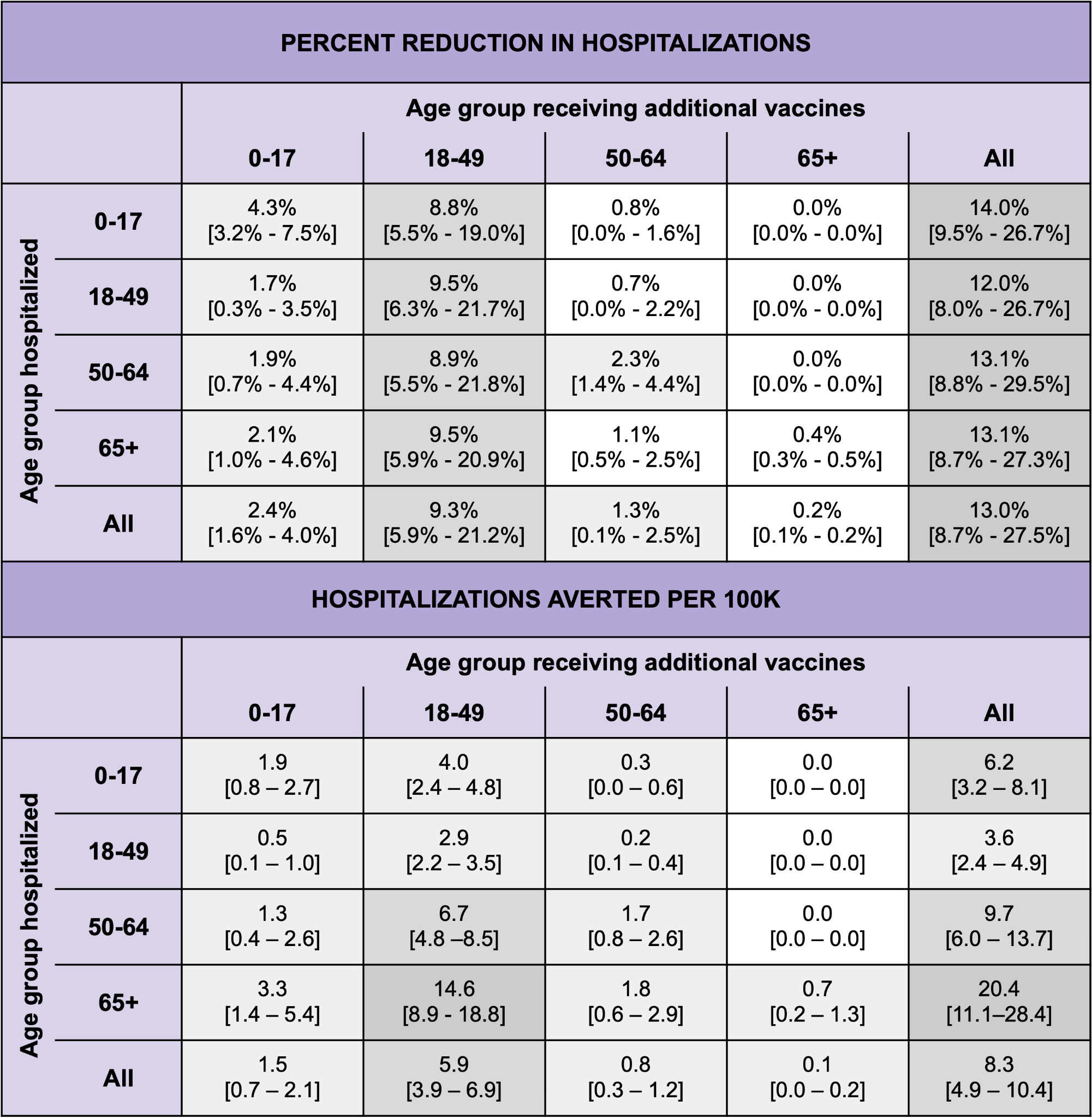

#### **A.4. Sensitivity analysis**

#### We conducted a sensitivity analysis by varying VE against hospitalized infection(1) and VE against infection(2) based on the upper and lower bounds of the 95% confidence intervals provided in **Table 1**. For each pair of values, we derived the corresponding VE against hospitalization given infection (**Table S.A.4**).

**Table S.A.4: Sensitivity analysis scenarios.** Each scenario assumes a different set of age-specific vaccine effectiveness (VE) against infection, hospitalized-infection, and hospitalization conditional upon infection during the 2022-2023 influenza season.

|  | **Age group (years)** | | | |
| --- | --- | --- | --- | --- |
|  | **0-17** | **18-49** | **50-64** | **65+** |
| **BASELINE SCENARIO** | | | | |
| **VE against infection** (2) | 43% | 21% | 21% | 0% |
| **VE against hospitalized infection** (3) | 68% | 23% | 23% | 41% |
| **VE against hospitalization given infection** (derived*) | 44% | 3% | 3% | 41% |
| **LOW EFFICACY SCENARIO** | | | | |
| **VE against infection** | 33% | 6% | 6% | 0% |
| **VE against hospitalized infection** | 46% | 10% | 10% | 33% |
| **VE against hospitalization given infection** (derived*) | 25% | 4% | 4% | 33% |
| **HIGH EFFICACY SCENARIO** | | | | |
| **VE against infection** | 55% | 34% | 34% | 14% |
| **VE against hospitalized infection** | 81% | 39% | 39% | 48% |
| **VE against hospitalization given infection** (derived*) | 58% | 8% | 8% | 40% |

* VE against hospitalization conditional upon infection is derived assuming independence as given by $VE_{hosp|inf}= 1-(1-VE_{hosp})/(1-VE_{inf})$.

We compared the estimated influenza hospitalizations averted (**Table S.A.5**) and additional influenza hospitalizations averted (**Table S.A.6**) across the different VE scenarios.

**Table S.A.5: Estimated influenza hospitalizations averted in each age group, across the three different vaccine efficacy scenarios.** Percent averted and averted per 100K are the estimated reductions in hospitalizations relative to the number estimated under a *no vaccination* scenario. Values are medians and 95% confidence intervals based on 1,000 pairs of stochastic simulations.

| **PERCENT REDUCTIONS IN HOSPITALIZATIONS** | | | |
| --- | --- | --- | --- |
|  | **Low VE scenario** | **Baseline Scenario** | **High VE scenario** |
| **0-17** | 12.3% [8.3%, 20.8%] | 26.1% [19.9%, 39.3%] | 34.9% [9.4%, 83.9%] |
| **18-49** | 6.8% [3.7%, 13.9%] | 14.8% [9.3%, 25.8%] | 23.7% [3.1%, 74.5%] |
| **50-64** | 7.8% [4.2%, 14.2%] | 18.3% [11.9%, 30.9%] | 25.6% [3.9%, 81.1%] |
| **65+** | 19.5% [16.3%, 24.5%] | 32.4% [27.5%, 41.1%] | 34.8% [7.2%, 72.8%] |
| **All** | 13.6% [10.7%, 18.8%] | 25.4% [20.5%, 35.3%] | 31.0% [9.6%, 78.1%] |
| **HOSPITALIZATIONS AVERTED PER 100K** | | | |
|  | **Low VE scenario** | **Baseline Scenario** | **High VE scenario** |
| **0-17** | 6.4 [2.9, 9.6] | 15.9 [8.7- 21.3] | 28.0 [0.0, 43.4] |
| **18-49** | 2.1 [1.2, 3.1] | 5.0 [3.1 - 6.9] | 8.0 [0.0, 11.1] |
| **50-64** | 6.2 [3.1, 8.5] | 17.3 [10.7 - 21.4] | 28.0 [0.0, 37.3] |
| **65+** | 38.8 [10.4, 75.6] | 80.4 [33.5 - 125.3] | 120.5 [0.2, 330.4] |
| **All** | 9.8 [3.5, 17.0] | 22.3 [11.6 - 31.4] | 37.2 [3.7, 72.0] |

**Table S.A.6: Estimated *additional* influenza hospitalizations averted if 70% of the US population had received 2022-2023 influenza vaccines, compared across the three vaccine efficacy scenarios.** Percent averted and averted per 100K are the estimated reductions in hospitalizations relative to the *baseline* 2022-2023 vaccination scenario. Values are medians and 95% confidence intervals based on 1,000 pairs of stochastic simulations.

| **PERCENT REDUCTIONS IN HOSPITALIZATIONS** | | | |
| --- | --- | --- | --- |
|  | **Low VE scenario** | **Baseline Scenario** | **High VE scenario** |
| **0-17** | 4.1% [2.3%, 7.8%] | 14.0% [9.5%, 26.7%] | 20.0% [0.3%, 78.9%] |
| **18-49** | 4.2% [2.3%, 6.6%] | 12.0% [8.0%, 26.7%] | 21.8% [-4.1%, 72.6%] |
| **50-64** | 3.5% [1.8%, 8.4%] | 13.1% [8.8%, 29.5%] | 20.8% [-1.3%, 72.6%] |
| **65+** | 3.6% [1.7%, 7.3%] | 13.1% [8.7%, 27.3%] | 16.6% [-1.3%, 72.6%] |
| **All** | 3.8% [2.1%, 7.0%] | 13.0% [8.7%, 27.5%] | 18.1% [0.0%, 68.3%] |
| **HOSPITALIZATIONS AVERTED PER 100K** | | | |
|  | **Low VE scenario** | **Baseline Scenario** | **High VE scenario** |
| **0-17** | 1.9 [0.8, 2.7] | 6.2 [3.2, 8.1] | 10.4 [0.0, 15.7] |
| **18-49** | 1.2 [0.5, 1.7] | 3.6 [2.4, 4.9] | 6.7 [0.0, 9.2] |
| **50-64** | 2.6 [1.2, 3.9] | 9.7 [6.0, 13.7] | 18.2 [0.0, 25.9] |
| **65+** | 5.7 [1.5, 8.5] | 20.4 [11.1, 28.4] | 38.8 [0.0, 62.0] |
| **All** | 2.4 [0.9, 3.2] | 8.3 [4.9, 10.4] | 15.3 [0.0, 22.1] |

#### **A.5. Correlates of state-level influenza hospitalizations in 2022-2023**

We conducted a multilinear regression relating influenza hospitalizations per 100K between 16.99 and 160.89 to age-specific influenza vaccination rates across all 50 US states (**Table S.A.7**).

**Table S.A.7: Linear regression analyses of state-level influenza hospitalizations on age-specific vaccination rates during the 2022-2023 influenza season.** Bolded rows indicate statistically significant variables.

| **Regression of influenza hospitalizations on overall vaccine coverage (US states)** | | | | | |
| --- | --- | --- | --- | --- | --- |
|  | Coefficient | 95% CI | Standard Error | T-value | P-value |
| Intercept | 132.07 | [71.34, 192.78] | 30.20 | 4.37 | 0.0001 |
| Overall coverage | -1.29 | [-2.61, 0.03] | 0.66 | -1.96 | 0.0553 |
| **Regression of influenza hospitalizations on vaccine coverage in 18-49 year olds (US states)** | | | | | |
|  | Coefficient | 95% CI | Standard Error | T-value | P-value |
| Intercept | 127.14 | [85.63, 168.67] | 20.64 | 6.16 | 0.000001 |
| **18-49y coverage** | **-1.62** | **[-2.85, -0.39]** | **0.61** | **-2.65** | **0.0107** |
| **Multi-regression of influenza hospitalizations on age-specific vaccine coverage (US states)** | | | | | |
|  | Coefficient | 95% CI | Standard Error | T-value | P-value |
| Intercept | 82.89 | [-22.61, 188.38] | 52.45 | 1.58 | 0.1205 |
| 0-17 coverage | -3.57 | [-7.47, 0.34] | 1.94 | -1.84 | 0.0723 |
| **18-49 coverage** | **-9.21** | **[-16.45, -1.95]** | **3.60** | **-2.56** | **0.0140** |
| 50-64 coverage | 0.03 | [-3.71, 3.76] | 1.85 | 0.01 | 0.9883 |
| 65+ coverage | -2.32 | [-6.95, 2.31] | 2.30 | -1.01 | 0.3180 |
| Overall coverage | 13.77 | [-6.14, 30.68] | 8.39 | 1.64 | 0.1081 |

#### **A.6. Relationship between estimated 2022-2023 burden averted and additional burden averted at the 70% vaccination target across US states**

**Figure S.A.1** compares the median estimates for burden averted by 2022-2023 influenza vaccines and additional burden averted had 70% of age groups been vaccinated. The estimated slope of the relationship is 0.0633 [95% CI: -0.0648, 0.705], but is not significant.

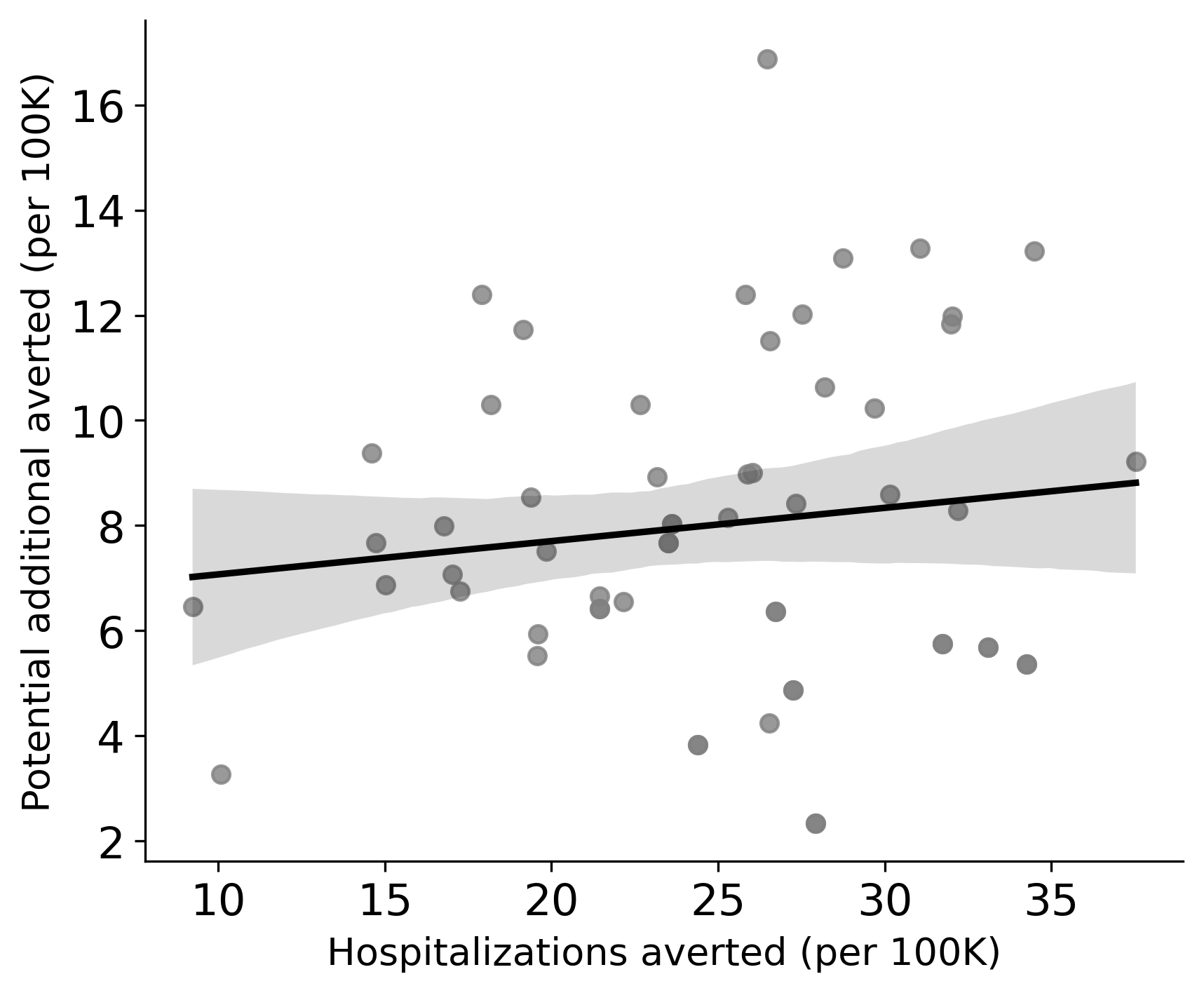

**Figure S.A.1**: **Estimated influenza hospitalizations averted by 2022-2023 vaccines compared to the projected additional hospitalizations averted had 70% of the US population received vaccines.** Each point corresponds to a US state; values are medians across 1,000 pairs of stochastic simulations. The line and shading represent the fitted regression line and 95% confidence band, respectively (*p*=0.3267).

### **Supplementary Material B: Modeling Details**

#### **B.1. Counterfactual Scenarios**

To retrospectively evaluate the impact of vaccination in the US during the 2022-2023 influenza season, we considered six types of counterfactual scenarios (Table S.B.1).

**Table S.B.1. Counterfactual scenarios.** All scenarios assume transmission rates obtained by fitting the baseline model to 2022-2023 influenza hospital admissions data.

| **Scenario Type** | **Description** |
| --- | --- |
| **Baseline** | Estimated 2022-2023 influenza vaccine coverage and estimated 2022-2023 VE against both infection and hospitalization, in all age groups. |
| **No vaccine** | No vaccination in the US. |
| **70% target** | 70% vaccine coverage in all age groups, with estimated 2022-2023 VE against both infection and hospitalization. |
| **Single age group - baseline** | Estimated 2022-2023 influenza vaccine coverage and estimated 2022-2023 VE against both infection and hospitalization, for a single age group. All other ages are not vaccinated. |
| **Single age group - 70%** | 70% vaccine coverage in a single age group and estimated 2022-2023 vaccine coverage in all other age groups, with estimated 2022-2023 VE against both infection and hospitalization. |
| **VE against infection only** | Estimated 2022-2023 influenza vaccination coverage and estimated 2022-2023 VE against infection, in all age groups. VE against hospitalization upon infection is set to zero in all age groups. |

**B2. Age-specific Counterfactual Scenarios**

To retrospectively evaluate the age-specific vaccination intervention in the US during the 2022-2023 season, we considered sixteen different age-specific scenarios (**Table S.B.2**). We estimated the age-specific hospitalizations averted by each age group with vaccinations (**Figure 1.C and Table S.A.2**) by comparing the Specific age group vac only scenario with the non-vaccine model. In addition, we estimated the age-specific hospitalizations additionally averted by each age group with 70% vaccination coverage by comparing the specific age group 70% uptake-only scenario with the baseline model (**Table S.A.3**).

**Table S.B.2. Sixteen counterfactual age-specific vaccine uptake scenarios for the 2022-2023 influenza season.**

|  | **SCENARIO TYPE** | | | |
| --- | --- | --- | --- | --- |
|  | **Singe group vaccination** | **Single group left out** | **Single group achieves 70% coverage** | **Single group left out of 70% coverage** |
| **0-17** | 0-17y group vaccinated at 2022-2023 levels  Other ages are unvaccinated | 0-17y group is unvaccinated  Other ages are vaccinated at 2022-2023 levels | 0-17y group achieves 70% coverage  Other ages are vaccinated at 2022-2023 levels | 0-17y group achieves vaccinated at 2022-2023 levels  Other ages achieve 70% coverage |
| **18-49** | 18-49y group vaccinated at 2022-2023 levels  Other ages are unvaccinated | 18-49y group is unvaccinated  Other ages are vaccinated at 2022-2023 levels | 18-49y group achieves 70% coverage  Other ages are vaccinated at 2022-2023 levels | 18-49y group achieves vaccinated at 2022-2023 levels  Other ages achieve 70% coverage |
| **50-64** | 50-64y group vaccinated at 2022-2023 levels  Other ages are unvaccinated | 50-64y group is unvaccinated  Other ages are vaccinated at 2022-2023 levels | 50-64y group achieves 70% coverage  Other ages are vaccinated at 2022-2023 levels | 50-64y group achieves vaccinated at 2022-2023 levels  Other ages achieve 70% coverage |
| **65+** | 65+y group vaccinated at 2022-2023 levels  Other ages are unvaccinated | 65+y group is unvaccinated  Other ages are vaccinated at 2022-2023 levels | 65+y group achieves 70% coverage  Other ages are vaccinated at 2022-2023 levels | 65+y group achieves vaccinated at 2022-2023 levels  Other ages achieve 70% coverage |

#### **B3. Mathematical Model Details**

We use an age-structured SEIRS model of influenza transmission, which explicitly tracks the immunity resulting from infection by either the influenza A(H1N1) or influenza A(H3N2) subtype and vaccination (**Figure 3**). The model tracks both *interseasonal immunity* that decreases rapidly at the onset of a new season and *intraseasonal immunity* that increases with new infections and vaccinations. For age group
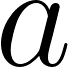
<https://www.codecogs.com/eqnedit.php?latex=a#0>, the changing levels of infection-derived protection against infection are given by:

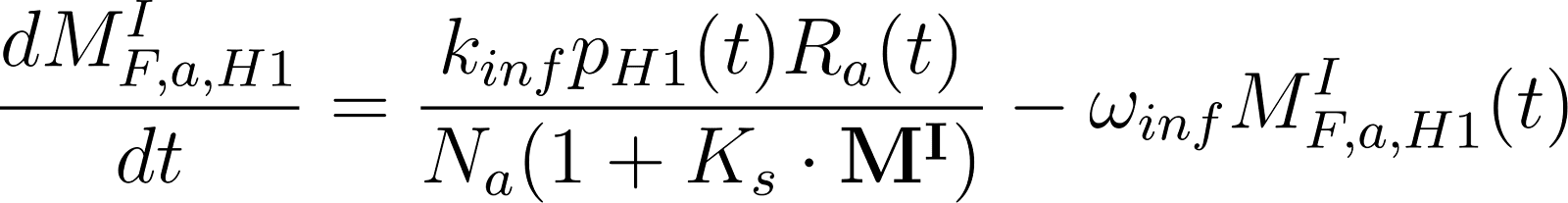
<https://www.codecogs.com/eqnedit.php?latex=%5Cfrac%7BdM_%7BF%2Ca%2CH1%7D%5EI%7D%7Bdt%7D%3D%5Cfrac%7Bk_%7Binf%7D%20p_%7BH1%7D(t)R_a(t)%7D%7BN_a(1%2BK_s%5Ccdot%20%5Cbf%7BM%5EI%7D)%7D-%5Comega_%7Binf%7DM_%7BF%2Ca%2CH1%7D%5EI(t)#0>

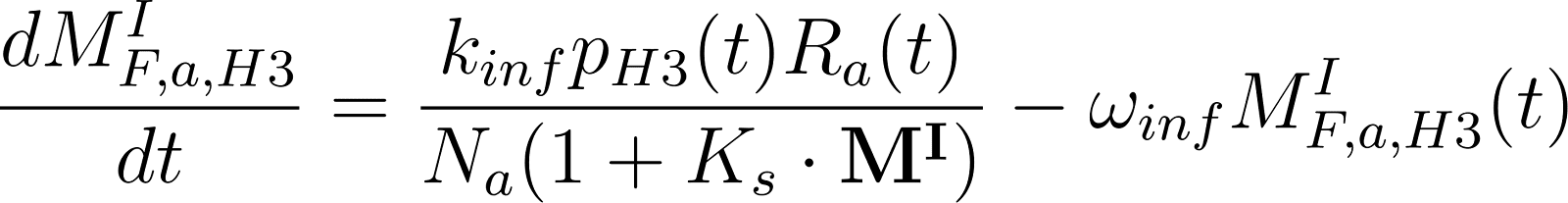
<https://www.codecogs.com/eqnedit.php?latex=%5Cfrac%7BdM_%7BF%2Ca%2CH3%7D%5EI%7D%7Bdt%7D%3D%5Cfrac%7Bk_%7Binf%7Dp_%7BH3%7D(t)R_a(t)%7D%7BN_a(1%2BK_s%5Ccdot%5Cbf%7BM%5EI%7D)%7D-%5Comega_%7Binf%7DM_%7BF%2Ca%2CH3%7D%5EI(t)#0>

where
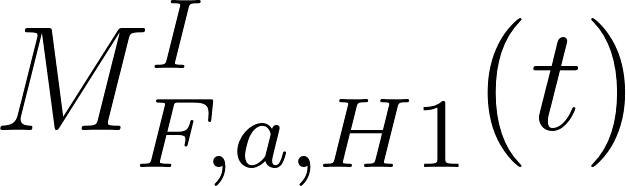
<https://www.codecogs.com/eqnedit.php?latex=M%5EI_%7BF%2Ca%2CH1%7D(t)#0> and
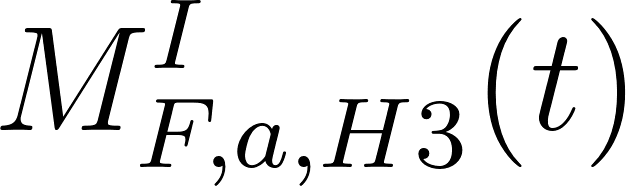
<https://www.codecogs.com/eqnedit.php?latex=M%5EI_%7BF%2Ca%2CH3%7D(t)#0> denote population-level immunity resulting from H1N1 infections and H3N2 infections, respectively, in age group
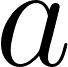
<https://www.codecogs.com/eqnedit.php?latex=a#0>.
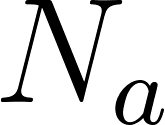
<https://www.codecogs.com/eqnedit.php?latex=N_a#0> is the total population of age group
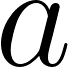
<https://www.codecogs.com/eqnedit.php?latex=a#0>,
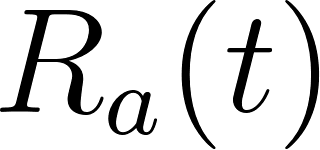
<https://www.codecogs.com/eqnedit.php?latex=R_a(t)#0> denotes the number of recovered individuals in age group
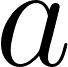
<https://www.codecogs.com/eqnedit.php?latex=a#0>.
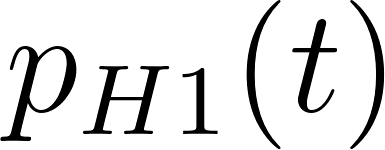
<https://www.codecogs.com/eqnedit.php?latex=p_%7BH1%7D(t)#0> and
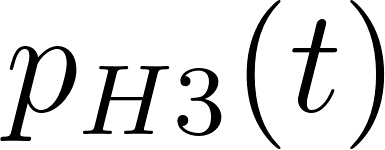
<https://www.codecogs.com/eqnedit.php?latex=p_%7BH3%7D(t)#0>denote the changing prevalence of H1N1 and H3N2 across all infections. However, since both the subtypes circulate in most seasons, these values are set to 1 and 0 throughout the season depending on the subtype dominant season, where the value 1 corresponds to prevalence of the dominant strain.

The model tracks the total immunity in the population, which increases by a factor of
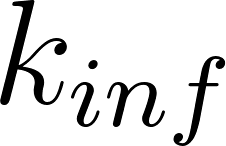
<https://www.codecogs.com/eqnedit.php?latex=k_%7Binf%7D#0> for each case that recovers and wanes at a rate of
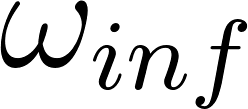
<https://www.codecogs.com/eqnedit.php?latex=%5Comega_%7Binf%7D#0>.
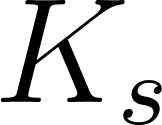
<https://www.codecogs.com/eqnedit.php?latex=K_s#0> is a positive constant modeling the saturation of antibody production in individuals who were previously infected. The changing levels of vaccine-derived protection against infection are given by:

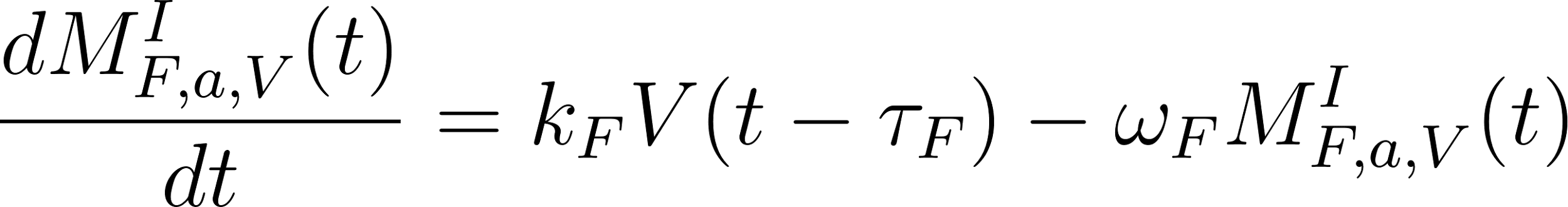
<https://www.codecogs.com/eqnedit.php?latex=%5Cfrac%7BdM_%7BF%2Ca%2CV%7D%5EI(t)%7D%7Bdt%7D%3Dk_%7BF%7DV(t-%5Ctau_F)-%5Comega_%7BF%7DM_%7BF%2Ca%2CV%7D%5EI(t)#0>

where
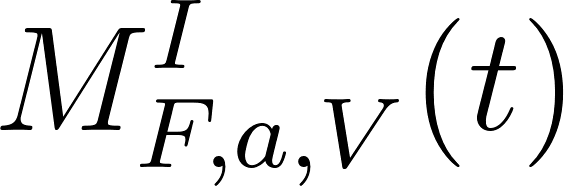
<https://www.codecogs.com/eqnedit.php?latex=M%5EI_%7BF%2Ca%2CV%7D(t)#0> represents population-level immunity derived from vaccines, respectively.
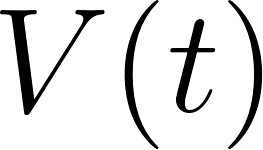
<https://www.codecogs.com/eqnedit.php?latex=V(t)#0> is the number of vaccine doses administered at time
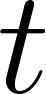
<https://www.codecogs.com/eqnedit.php?latex=t#0> and
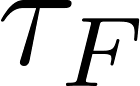
<https://www.codecogs.com/eqnedit.php?latex=%5Ctau_F#0> represents the delay in the number of days of dose administration. The total immunity in the population increases by a factor of
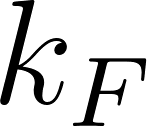
<https://www.codecogs.com/eqnedit.php?latex=k_F#0> for each dose of vaccine administered and then wanes at rates of
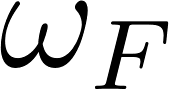
<https://www.codecogs.com/eqnedit.php?latex=%5Comega_F#0>. Similarly, we describe the changing levels of population immunity against hospitalization as given by:

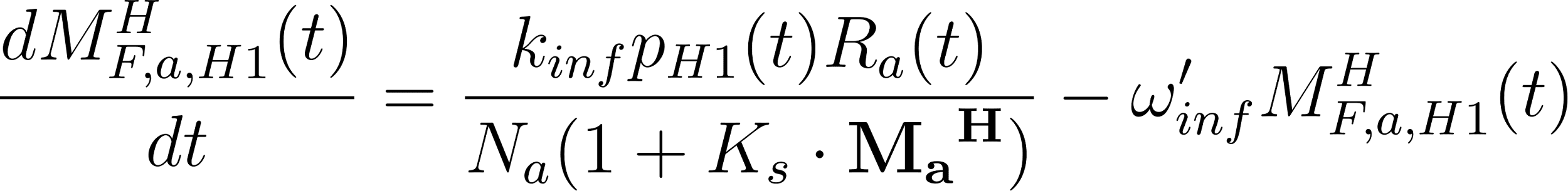
<https://www.codecogs.com/eqnedit.php?latex=%5Cfrac%7BdM_%7BF%2Ca%2CH1%7D%5EH(t)%7D%7Bdt%7D%3D%5Cfrac%7Bk_%7Binf%7Dp_%7BH1%7D(t)R_a(t)%7D%7BN_a(1%2BK_s%5Ccdot%5Cbf%7B%7BM_a%7D%5EH%7D)%7D-%5Comega_%7Binf%7D'M_%7BF%2Ca%2CH1%7D%5EH(t)#0>

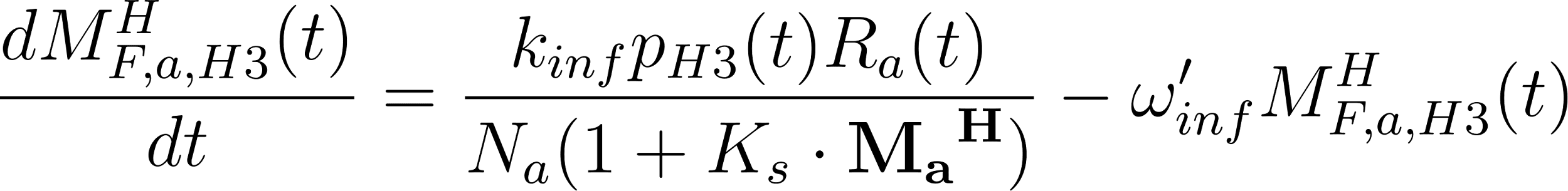
<https://www.codecogs.com/eqnedit.php?latex=%5Cfrac%7BdM_%7BF%2Ca%2CH3%7D%5EH(t)%7D%7Bdt%7D%3D%5Cfrac%7Bk_%7Binf%7Dp_%7BH3%7D(t)R_a(t)%7D%7BN_a(1%2BK_s%5Ccdot%5Cbf%7B%7BM_a%7D%5EH%7D)%7D-%5Comega_%7Binf%7D'M_%7BF%2Ca%2CH3%7D%5EH(t)#0>

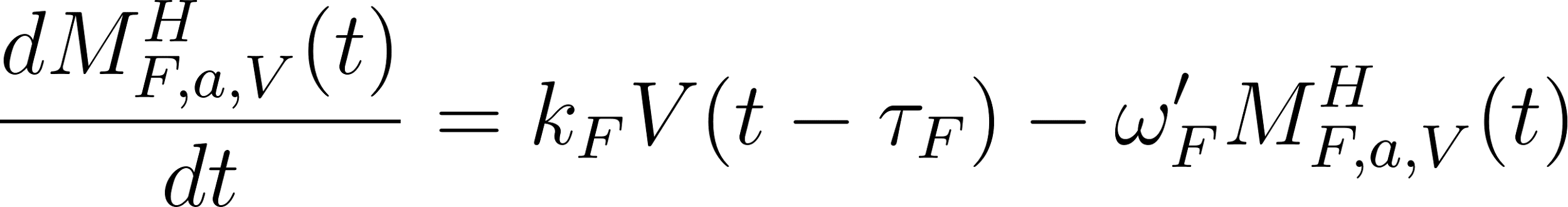
<https://www.codecogs.com/eqnedit.php?latex=%5Cfrac%7BdM_%7BF%2Ca%2CV%7D%5EH(t)%7D%7Bdt%7D%3Dk_%7BF%7DV(t-%5Ctau_F)-%5Comega_%7BF%7D'M_%7BF%2Ca%2CV%7D%5EH(t)#0>

where
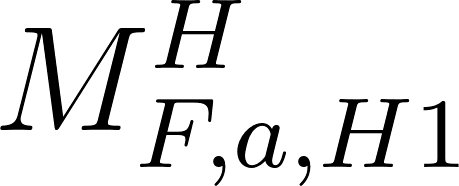
<https://www.codecogs.com/eqnedit.php?latex=M%5EH_%7BF%2Ca%2CH1%7D#0>*,*
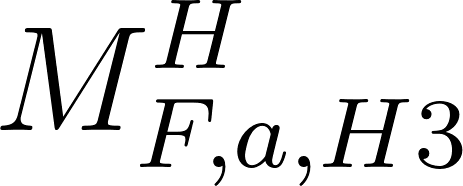
<https://www.codecogs.com/eqnedit.php?latex=M%5EH_%7BF%2Ca%2CH3%7D#0> and
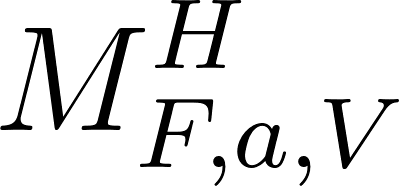
<https://www.codecogs.com/eqnedit.php?latex=M%5EH_%7BF%2Ca%2CV%7D#0> represent population-level immunity against influenza hospitalizations derived from infections by the H1N1 subtype, infections by the H3N2 subtype, and vaccines, respectively.
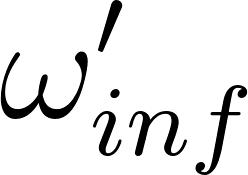
<https://www.codecogs.com/eqnedit.php?latex=%5Comega_%7Binf%7D'#0> denotes the rate of waning of population-level immunity against hospitalization and
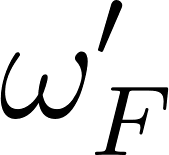
<https://www.codecogs.com/eqnedit.php?latex=%5Comega_F'#0> denotes the rate at which the immunity wanes for vaccine dose administered. These immunity variables modify the age-specific transitions among disease compartments as given by:

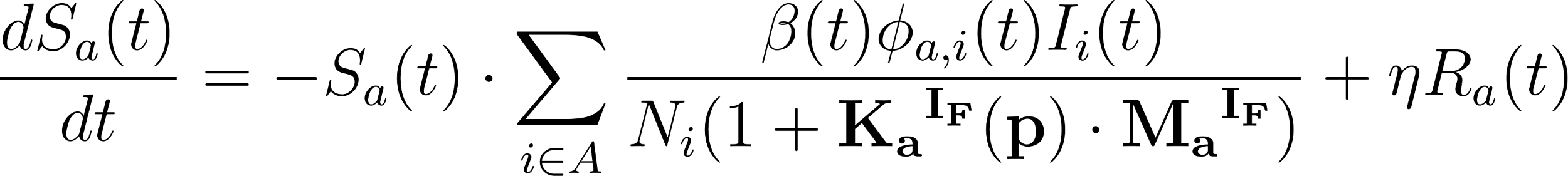
<https://www.codecogs.com/eqnedit.php?latex=%5Cfrac%7BdS_a(t)%7D%7Bdt%7D%3D-S_a(t)%5Ccdot%5Csum_%7Bi%5Cin%20A%7D%5E%7B%20%7D%5Cfrac%7B%5Cbeta(t)%5Cphi_%7Ba%2Ci%7D(t)I_i(t)%7D%7BN_i(1%2B%5Cbf%7B%7BK_a%7D%5E%7BI_F%7D(p)%7D%5Ccdot%5Cbf%7B%7BM_a%7D%5E%7BI_F%7D%7D)%7D%2B%5Ceta%20R_a(t)#0>

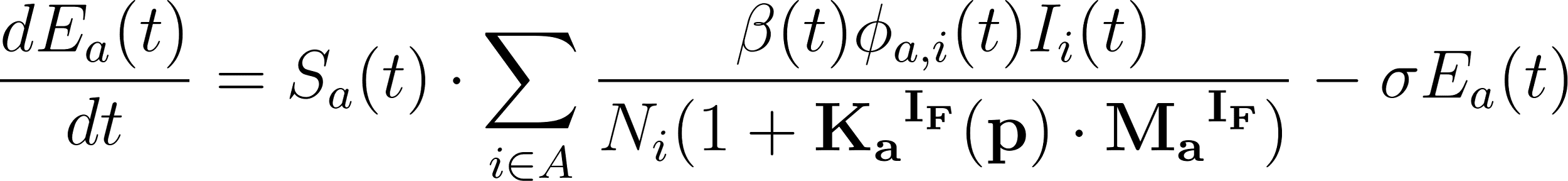
<https://www.codecogs.com/eqnedit.php?latex=%5Cfrac%7BdE_a(t)%7D%7Bdt%7D%3DS_a(t)%5Ccdot%5Csum_%7Bi%5Cin%20A%7D%5E%7B%20%7D%5Cfrac%7B%5Cbeta(t)%5Cphi_%7Ba%2Ci%7D(t)I_i(t)%7D%7BN_i(1%2B%5Cbf%7B%7BK_a%7D%5E%7BI_F%7D(p)%7D%5Ccdot%5Cbf%7B%7BM_a%7D%5E%7BI_F%7D%7D)%7D-%5Csigma%20E_a(t)#0>

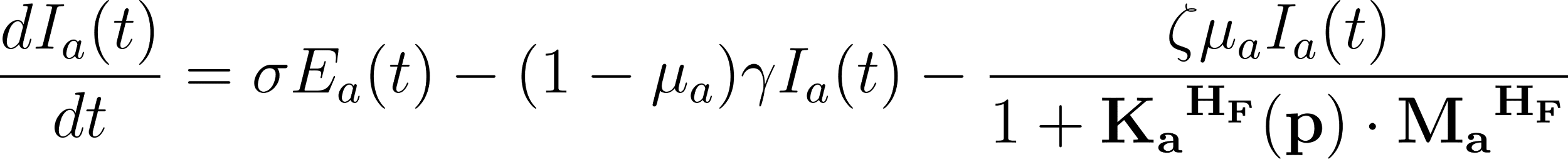
<https://www.codecogs.com/eqnedit.php?latex=%5Cfrac%7BdI_a(t)%7D%7Bdt%7D%3D%5Csigma%20E_a(t)-(1-%5Cmu_a)%5Cgamma%20I_a(t)-%5Cfrac%7B%5Czeta%5Cmu_a%20I_a(t)%7D%7B1%2B%5Cbf%7B%7BK_a%7D%5E%7BH_F%7D(p)%7D%5Ccdot%5Cbf%7B%7BM_a%7D%5E%7BH_F%7D%7D%7D#0>

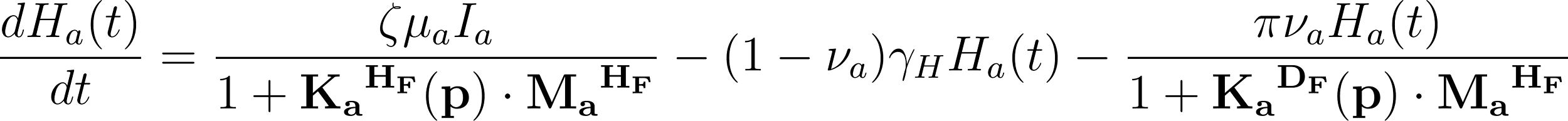
<https://www.codecogs.com/eqnedit.php?latex=%5Cfrac%7BdH_a(t)%7D%7Bdt%7D%3D%5Cfrac%7B%5Czeta%5Cmu_a%20I_a%7D%7B1%2B%5Cbf%7B%7BK_a%7D%5E%7BH_F%7D(p)%7D%5Ccdot%5Cbf%7B%7BM_a%7D%5E%7BH_F%7D%7D%7D-(1-%5Cnu_a)%5Cgamma_HH_a(t)-%5Cfrac%7B%5Cpi%5Cnu_aH_a(t)%7D%7B1%2B%5Cbf%7B%7BK_a%7D%5E%7BD_F%7D(p)%7D%5Ccdot%5Cbf%7B%7BM_a%7D%5E%7BH_F%7D%7D%7D#0>

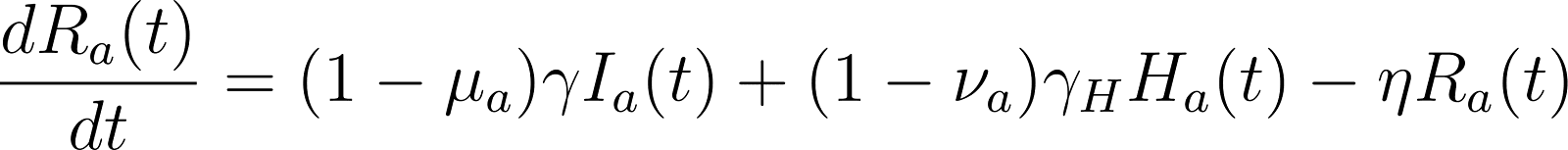
<https://www.codecogs.com/eqnedit.php?latex=%5Cfrac%7BdR_a(t)%7D%7Bdt%7D%3D(1-%5Cmu_a)%5Cgamma%20I_a(t)%20%2B(1-%5Cnu_a)%5Cgamma_H%20H_a(t)%20-%5Ceta%20R_a(t)#0>

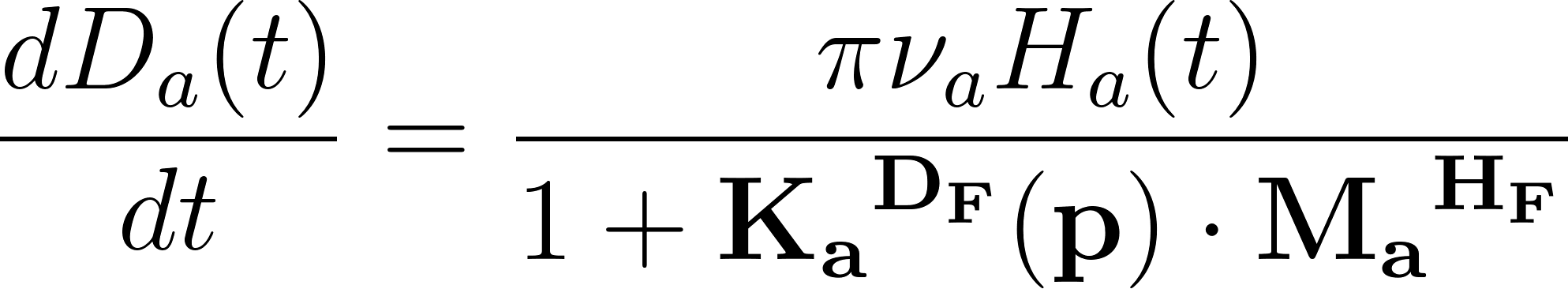
<https://www.codecogs.com/eqnedit.php?latex=%5Cfrac%7BdD_a(t)%7D%7Bdt%7D%3D%5Cfrac%7B%5Cpi%5Cnu_aH_a(t)%7D%7B1%2B%5Cbf%7B%7BK_a%7D%5E%7BD_F%7D(p)%7D%5Ccdot%5Cbf%7B%7BM_a%7D%5E%7BH_F%7D%7D%7D#0>

where
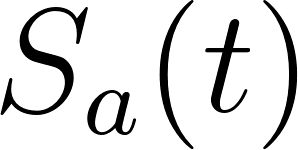
<https://www.codecogs.com/eqnedit.php?latex=S_a(t)#0>,
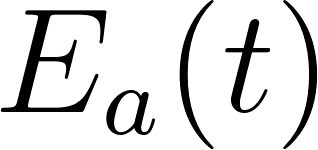
<https://www.codecogs.com/eqnedit.php?latex=E_a(t)#0>,
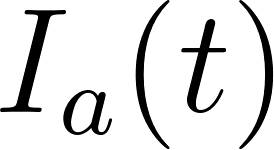
<https://www.codecogs.com/eqnedit.php?latex=I_a(t)#0>,
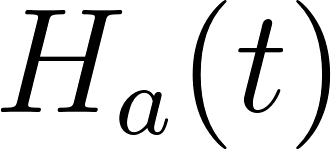
<https://www.codecogs.com/eqnedit.php?latex=H_a(t)#0> and
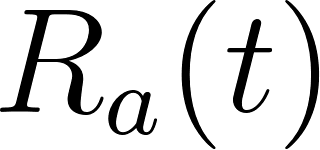
<https://www.codecogs.com/eqnedit.php?latex=R_a(t)#0> are age-specific numbers of people who are in the susceptible, exposed, infectious, hospitalized, recovered and death compartments, respectively, at time
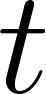
<https://www.codecogs.com/eqnedit.php?latex=t#0>. The time-dependent transmission rate is given by
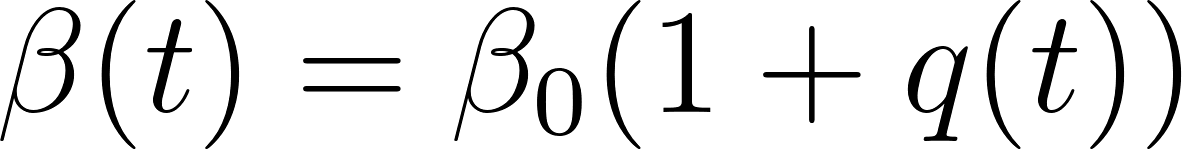
<https://www.codecogs.com/eqnedit.php?latex=%5Cbeta(t)%3D%5Cbeta_0%20(1%2Bq(t))#0>, where
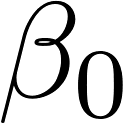
<https://www.codecogs.com/eqnedit.php?latex=%5Cbeta_0#0> is the transmission rate and
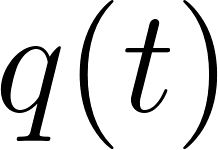
<https://www.codecogs.com/eqnedit.php?latex=q(t)#0> is a seasonality parameter based on absolute humidity (see below). The mixing rates between age groups
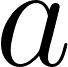
<https://www.codecogs.com/eqnedit.php?latex=a#0> and
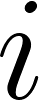
<https://www.codecogs.com/eqnedit.php?latex=i#0>,
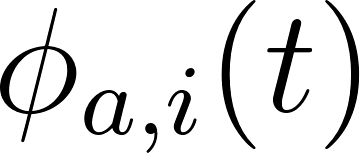
<https://www.codecogs.com/eqnedit.php?latex=%5Cphi_%7Ba%2Ci%7D(t)#0>, are based on published contact matrices (see below). The transition parameters
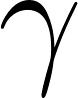
<https://www.codecogs.com/eqnedit.php?latex=%5Cgamma#0>,
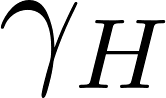
<https://www.codecogs.com/eqnedit.php?latex=%5Cgamma_H#0> denote the recovery rates for the
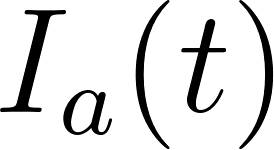
<https://www.codecogs.com/eqnedit.php?latex=I_a(t)#0> and
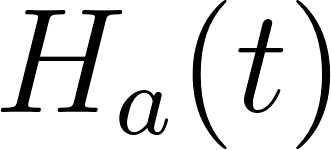
<https://www.codecogs.com/eqnedit.php?latex=H_a(t)#0> compartments, respectively,
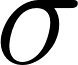
<https://www.codecogs.com/eqnedit.php?latex=%5Csigma#0> denotes the transition rate out of exposed compartment,
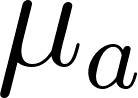
<https://www.codecogs.com/eqnedit.php?latex=%5Cmu_a#0> is the infection hospitalization rate,
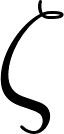
<https://www.codecogs.com/eqnedit.php?latex=%5Czeta#0> is the transition rate from
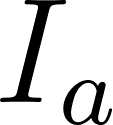
<https://www.codecogs.com/eqnedit.php?latex=I_a#0> to
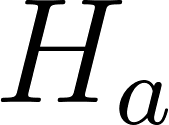
<https://www.codecogs.com/eqnedit.php?latex=H_a#0>,
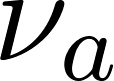
<https://www.codecogs.com/eqnedit.php?latex=%5Cnu_a#0> is the in-hospital mortality rate,
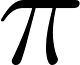
<https://www.codecogs.com/eqnedit.php?latex=%5Cpi#0> is the transition rate from
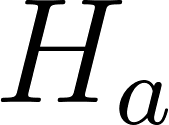
<https://www.codecogs.com/eqnedit.php?latex=H_a#0> to
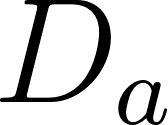
<https://www.codecogs.com/eqnedit.php?latex=D_a#0>, and
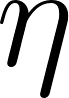
<https://www.codecogs.com/eqnedit.php?latex=%5Ceta#0> is the rate at which recovered individuals become susceptible again.
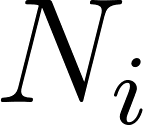
<https://www.codecogs.com/eqnedit.php?latex=N_i#0> is the total population of the age group
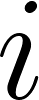
<https://www.codecogs.com/eqnedit.php?latex=i#0>. The vectors
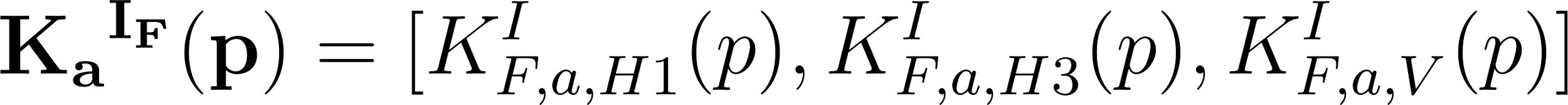
<https://www.codecogs.com/eqnedit.php?latex=%7B%5Cbf%7B%7BK_a%7D%5E%7BI_F%7D%20(p)%7D%7D%20%3D%20%5B%20K%5EI_%7BF%2Ca%2CH1%7D(p)%2CK%5EI_%7BF%2Ca%2CH3%7D(p)%2CK%5EI_%7BF%2Ca%2CV%7D(p)%5D#0>,
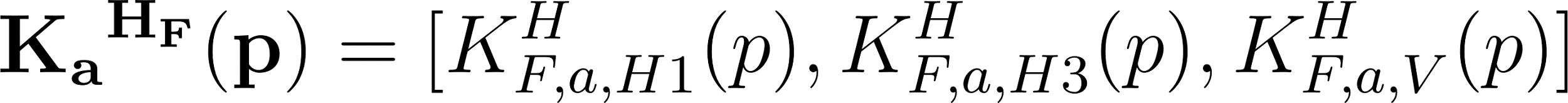
<https://www.codecogs.com/eqnedit.php?latex=%7B%5Cbf%7B%7BK_a%7D%5E%7BH_F%7D%20(p)%7D%7D%20%3D%20%5B%20K%5EH_%7BF%2Ca%2CH1%7D(p)%2CK%5EH_%7BF%2Ca%2CH3%7D(p)%2CK%5EH_%7BF%2Ca%2CV%7D(p)%5D#0> and
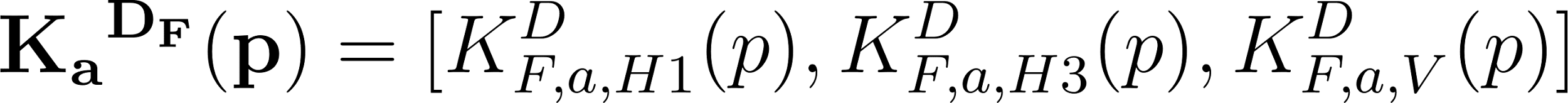
<https://www.codecogs.com/eqnedit.php?latex=%7B%5Cbf%7B%7BK_a%7D%5E%7BD_F%7D%20(p)%7D%7D%20%3D%20%5B%20K%5ED_%7BF%2Ca%2CH1%7D(p)%2CK%5ED_%7BF%2Ca%2CH3%7D(p)%2CK%5ED_%7BF%2Ca%2CV%7D(p)%5D#0> contain positive constants that describe efficacy of immunity in reducing the rate of influenza infection, hospitalization, and death, respectively. As described above,
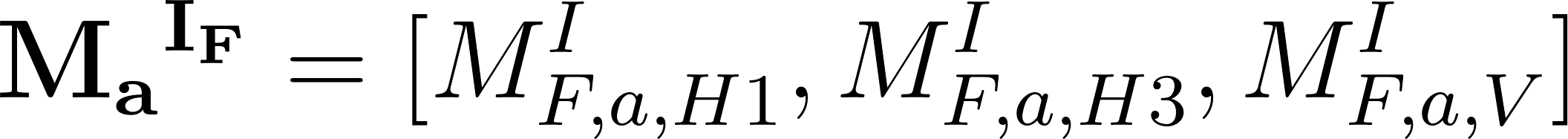
<https://www.codecogs.com/eqnedit.php?latex=%7B%5Cbf%7B%7BM_a%7D%5E%7BI_F%7D%7D%7D%20%3D%20%5B%20M%5EI_%7BF%2Ca%2CH1%7D%2CM%5EI_%7BF%2Ca%2CH3%7D%2CM%5EI_%7BF%2Ca%2CV%7D%5D#0> and
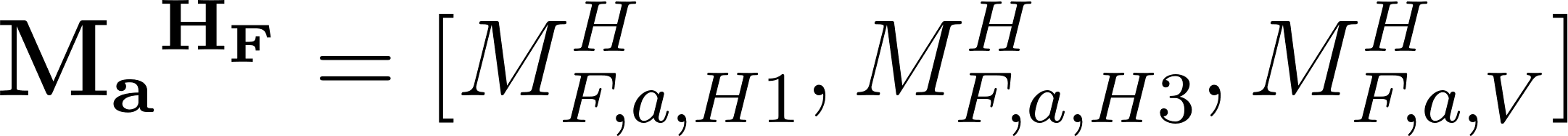
<https://www.codecogs.com/eqnedit.php?latex=%7B%5Cbf%7B%7BM_a%7D%5E%7BH_F%7D%7D%7D%20%3D%20%5B%20M%5EH_%7BF%2Ca%2CH1%7D%2CM%5EH_%7BF%2Ca%2CH3%7D%2CM%5EH_%7BF%2Ca%2CV%7D%5D#0> are two vectors consisting of state variables that describe the protection levels derived from natural infection and vaccination against infection and hospitalization, respectively. The overall structure of the model is similar to that of the COVID-19 model, with the exception of influenza subtypes, vaccination, and seasonality features. The details of the Influenza model parameter values are summarized in **Table S.B.3**.

The model was previously developed, validated, and applied to provide projections for the US Influenza Scenario Modeling Hub, 2022/2023 Round 1 to 3, and 2023/2024 Round 1(4).

#### **B4. Seasonality**

We incorporate humidity-based seasonality in our influenza and RSV models(5). We collected the absolute humidity data from 2006 to 2020 at the National Oceanic and Atmospheric Administration (NOAA)(6). We estimated the daily absolute humidity by averaging the collected data for the corresponding dates. We incorporate a seasonality parameter *q(t)* into the model equations as follows:

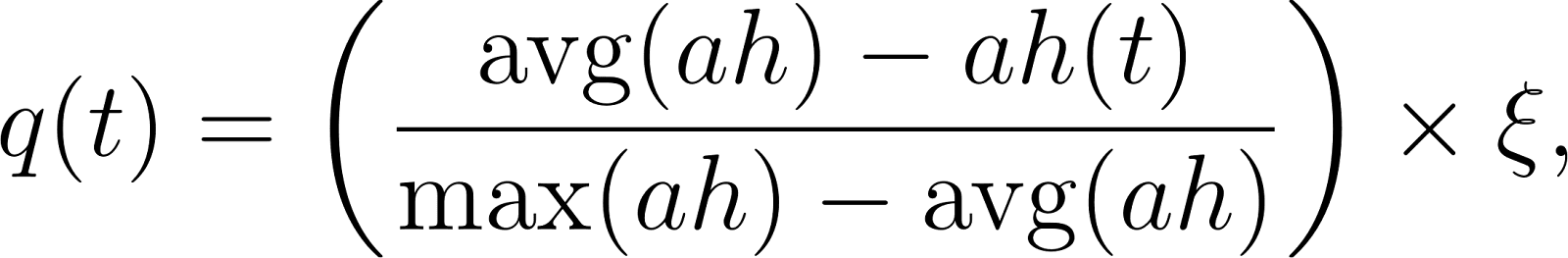
<https://www.codecogs.com/eqnedit.php?latex=%20q(t)%20%3D%20%5Cleft(%5Cfrac%7B%5Ctext%7Bavg%7D(ah)-ah(t)%7D%7B%5Cmax(ah)-%5Ctext%7Bavg%7D(ah)%7D%5Cright)%5Ctimes%20%5Cxi%2C#0>

where
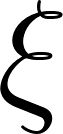
<https://www.codecogs.com/eqnedit.php?latex=%5Cxi#0> is the magnitude of the seasonality impact on transmissibility and
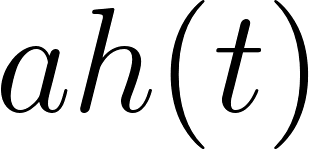
<https://www.codecogs.com/eqnedit.php?latex=ah(t)#0> is the estimated absolute humidity on a calendar day
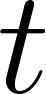
<https://www.codecogs.com/eqnedit.php?latex=t#0> (based on the fitted cosine model),
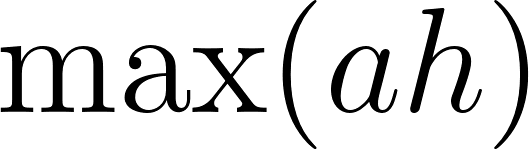
<https://www.codecogs.com/eqnedit.php?latex=%5Cmax(ah)#0>is the value of the maximum absolute humidity in a year, and
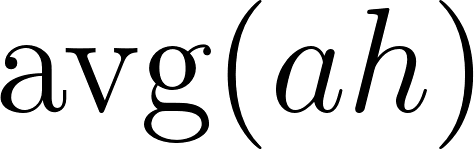
<https://www.codecogs.com/eqnedit.php?latex=%5Ctext%7Bavg%7D(ah)#0> is the value of the average maximum absolute humidity in a year.

#### **B5. Behavior and Contact Matrix**

We model daily age-specific mixing patterns using published estimates for venue-specific (all locations, school, and work) contact rates in the US(7). We assume that schools close on all weekends, during a two week winter break (December 18 to January 02), and throughout the summer break (June-August). Workplaces are assumed to be closed during the weekends. The overall contact matrix on day *t* is given by:

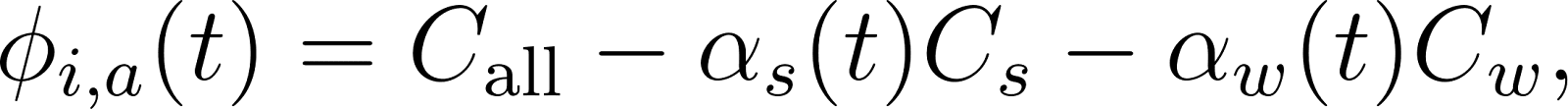
<https://www.codecogs.com/eqnedit.php?latex=%5Cphi_%7Bi%2Ca%7D(t)%20%3D%20C_%7B%5Ctext%7Ball%7D%7D%20-%20%5Calpha_s(t)%20C_s%20-%20%5Calpha_w(t)%20C_w%2C%20#0>

where
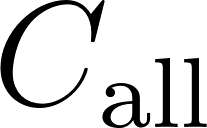
<https://www.codecogs.com/eqnedit.php?latex=C_%7B%5Ctext%7Ball%7D%7D#0>*,*
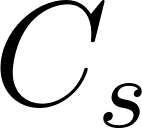
<https://www.codecogs.com/eqnedit.php?latex=C_%7Bs%7D#0> *and*
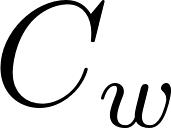
<https://www.codecogs.com/eqnedit.php?latex=C_%7Bw%7D#0> are the estimated age-specific contact matrices for all locations, schools, and workplaces, respectively.
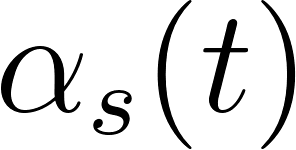
<https://www.codecogs.com/eqnedit.php?latex=%5Calpha_%7Bs%7D(t)#0> *and*
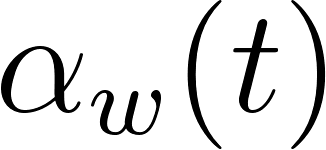
<https://www.codecogs.com/eqnedit.php?latex=%5Calpha_%7Bw%7D(t)#0> are time-dependent functions that describe the opening or closure of schools and workplaces, they equal 0 if the location is closed and 1 if it is closed. For the SARS-CoV-2 and influenza models, we consider six age groups [0-4, 5-11, 12-18, 19-49, 50-64, 65+] and assume the following contact matrices:

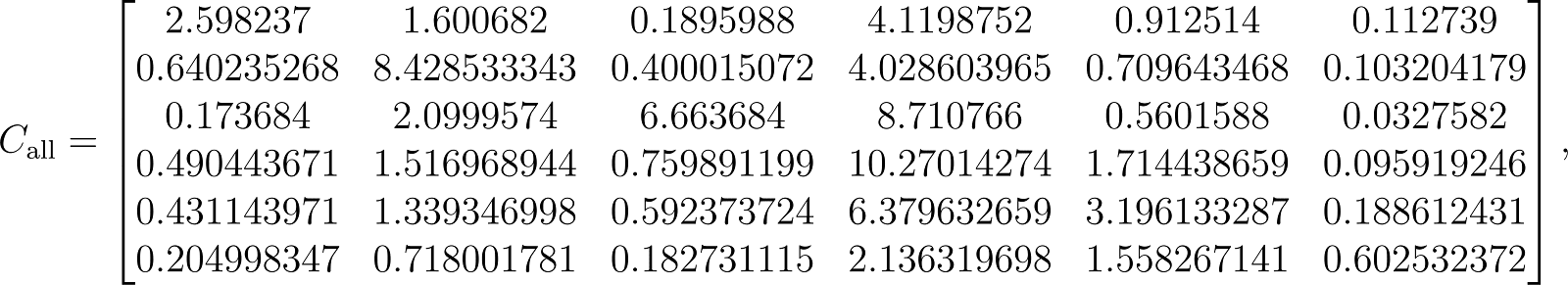
<https://www.codecogs.com/eqnedit.php?latex=C_%7B%5Ctext%7Ball%7D%7D%3D%5Cbegin%7Bbmatrix%7D%202.598237%20%20%26%201.600682%20%26%200.1895988%20%20%26%20%204.1198752%20%20%26%200.912514%20%26%20%200.112739%20%5C%5C%5C%5C%200.640235268%20%20%26%20%208.428533343%20%20%26%20%200.400015072%20%26%20%20%204.028603965%20%20%26%20%200.709643468%20%26%20%20%20%200.103204179%20%20%5C%5C%5C%5C%200.173684%20%20%26%20%202.0999574%20%20%26%20%206.663684%20%26%20%20%208.710766%20%26%20%20%20%200.5601588%20%26%20%20%200.0327582%20%5C%5C%5C%5C%200.490443671%20%20%26%20%201.516968944%20%26%20%20%200.759891199%20%20%26%20%2010.27014274%20%20%20%20%26%201.714438659%20%20%26%20%200.095919246%20%5C%5C%5C%5C%200.431143971%20%20%26%20%201.339346998%20%26%20%20%200.592373724%20%20%20%26%206.379632659%20%26%20%20%203.196133287%20%20%26%20%200.188612431%20%5C%5C%5C%5C%200.204998347%20%20%26%20%200.718001781%20%26%20%20%200.182731115%20%20%26%20%202.136319698%20%20%26%20%201.558267141%20%26%20%20%200.602532372%20%5Cend%7Bbmatrix%7D%2C#0>

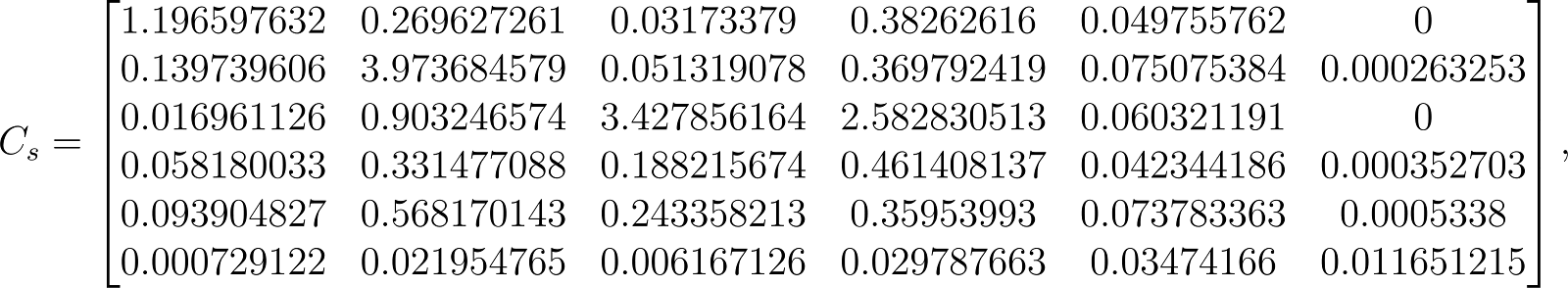
<https://www.codecogs.com/eqnedit.php?latex=C_%7Bs%7D%3D%5Cbegin%7Bbmatrix%7D%201.196597632%20%20%26%20%200.269627261%20%26%20%20%20%200.03173379%20%26%20%20%200.38262616%20%26%20%20%200.049755762%20%20%26%20%200%20%5C%5C%5C%5C%200.139739606%20%20%26%20%203.973684579%20%20%26%20%200.051319078%20%20%26%20%200.369792419%20%20%26%20%200.075075384%20%20%26%20%200.000263253%20%20%5C%5C%5C%5C%200.016961126%20%20%26%20%200.903246574%20%20%20%26%203.427856164%20%20%20%20%26%202.582830513%20%20%26%20%20%200.060321191%20%26%20%20%200%20%5C%5C%5C%5C%200.058180033%20%20%26%20%200.331477088%20%26%20%20%200.188215674%20%20%26%20%200.461408137%20%26%20%20%200.042344186%20%20%26%20%200.000352703%20%5C%5C%5C%5C%200.093904827%20%20%26%20%200.568170143%20%20%20%26%200.243358213%20%20%26%20%200.35953993%20%20%20%26%200.073783363%20%20%26%20%200.0005338%20%5C%5C%5C%5C%200.000729122%20%20%26%20%200.021954765%20%20%26%20%200.006167126%20%20%26%20%200.029787663%20%26%20%20%200.03474166%20%20%26%20%200.011651215%20%5Cend%7Bbmatrix%7D%2C#0>

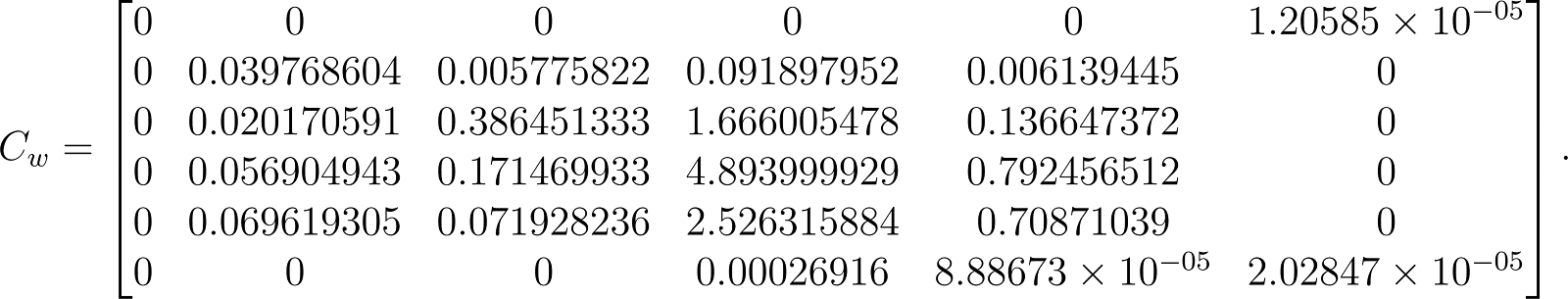
<https://www.codecogs.com/eqnedit.php?latex=C_%7Bw%7D%3D%5Cbegin%7Bbmatrix%7D%200%20%20%26%20%200%20%26%20%20%200%20%26%20%20%200%20%20%26%20%200%20%20%26%20%201.20585%20%5Ctimes%2010%5E%7B-05%7D%20%5C%5C%5C%5C%200%20%20%26%20%200.039768604%20%20%26%20%200.005775822%20%20%26%20%200.091897952%20%20%26%20%200.006139445%20%20%26%20%200%20%20%5C%5C%5C%5C%200%20%20%26%20%200.020170591%20%20%26%20%200.386451333%20%20%20%20%26%201.666005478%20%20%26%20%200.136647372%20%20%26%20%200%20%5C%5C0%20%20%20%26%200.056904943%20%20%26%20%200.171469933%20%26%20%20%204.893999929%20%20%20%20%26%200.792456512%20%20%26%20%20%200%20%5C%5C%5C%5C%200%20%20%20%26%200.069619305%20%20%26%20%200.071928236%20%20%26%20%202.526315884%20%20%26%20%200.70871039%20%20%20%20%26%200%20%20%5C%5C%5C%5C%200%20%26%20%200%20%26%20%20%20%200%20%26%20%20%20%200.00026916%20%26%20%20%208.88673%5Ctimes%2010%5E%7B-05%7D%20%20%20%26%20%202.02847%5Ctimes%2010%5E%7B-05%7D%20%5Cend%7Bbmatrix%7D.#0>

#### **B6. Stochasticity**

We incorporate stochasticity into our model by both varying the daily transmission rate
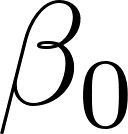
<https://www.codecogs.com/eqnedit.php?latex=%5Cbeta_0#0> and introducing variation into the transition rates between disease compartments. For each day, we draw a random value from the estimated normal distribution of
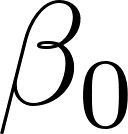
<https://www.codecogs.com/eqnedit.php?latex=%5Cbeta_0#0> values (**Table S.B.3**). We introduce stochasticity into the disease progression models using the tau-leap method(8), in which the transition rates are sampled from Poisson distribution with a mean equal to the deterministic rates multiplied by the time step
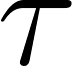
<https://www.codecogs.com/eqnedit.php?latex=%5Ctau#0>.

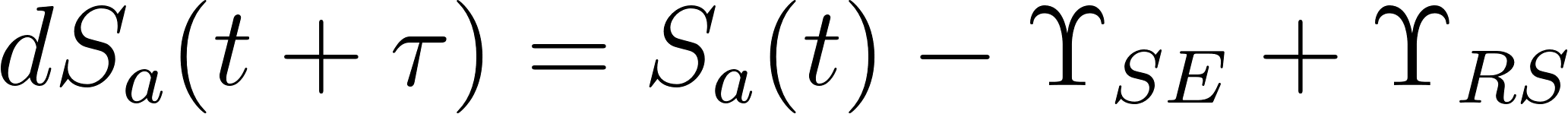
<https://www.codecogs.com/eqnedit.php?latex=%7BdS_a(t%2B%5Ctau)%7D%20%3D%20S_a(t)%20-%20%5CUpsilon_%7BSE%7D%20%2B%20%5CUpsilon_%7BRS%7D#0>

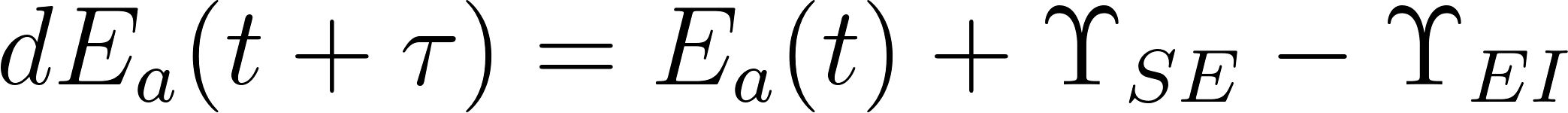
<https://www.codecogs.com/eqnedit.php?latex=%7BdE_a(t%2B%5Ctau)%7D%20%3D%20E_a(t)%20%2B%20%5CUpsilon_%7BSE%7D%20-%20%5CUpsilon_%7BEI%7D#0>

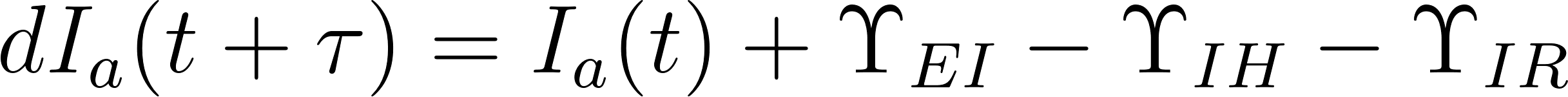
<https://www.codecogs.com/eqnedit.php?latex=%7BdI_a(t%2B%5Ctau)%7D%20%3D%20I_a(t)%20%2B%20%5CUpsilon_%7BEI%7D%20-%20%5CUpsilon_%7BIH%7D%20-%20%5CUpsilon_%7BIR%7D#0>

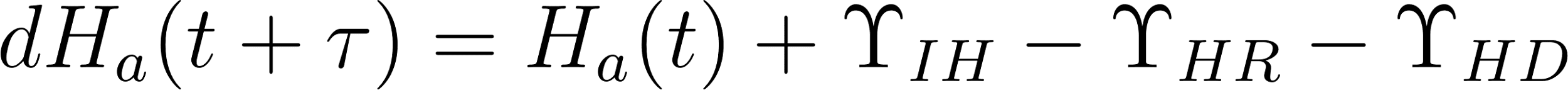
<https://www.codecogs.com/eqnedit.php?latex=%7BdH_a(t%2B%5Ctau)%7D%20%3D%20H_a(t)%20%2B%20%5CUpsilon_%7BIH%7D%20-%20%5CUpsilon_%7BHR%7D%20-%20%5CUpsilon_%7BHD%7D#0>

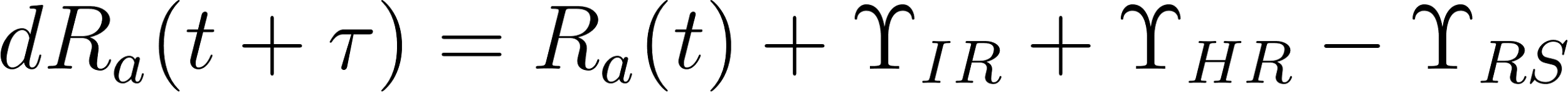
<https://www.codecogs.com/eqnedit.php?latex=%7BdR_a(t%2B%5Ctau)%7D%20%3D%20R_a(t)%20%2B%20%5CUpsilon_%7BIR%7D%20%2B%20%5CUpsilon_%7BHR%7D%20-%20%5CUpsilon_%7BRS%7D#0>

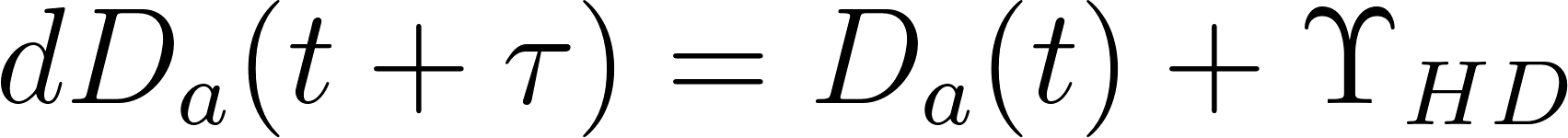
<https://www.codecogs.com/eqnedit.php?latex=%7BdD_a(t%2B%5Ctau)%7D%20%3D%20D_a(t)%20%2B%20%5CUpsilon_%7BHD%7D#0>

where the transition between compartments is as below:

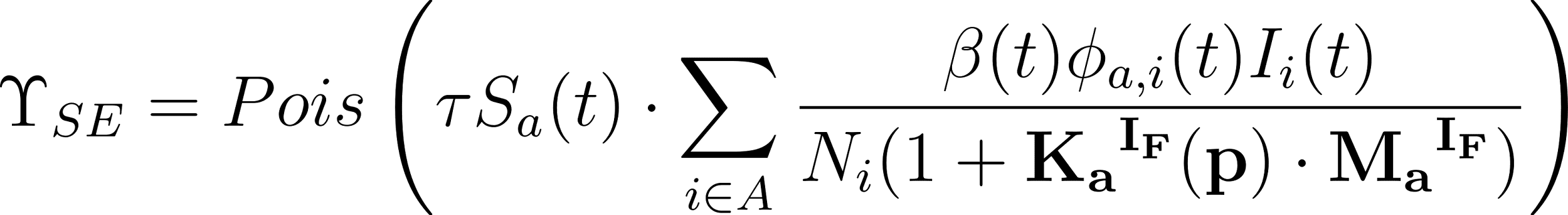
<https://www.codecogs.com/eqnedit.php?latex=%20%5CUpsilon_%7BSE%7D%20%3D%20Pois%5Cleft(%5Ctau%20S_a(t)%5Ccdot%5Csum_%7Bi%5Cin%20A%7D%5E%7B%20%7D%5Cfrac%7B%5Cbeta(t)%5Cphi_%7Ba%2Ci%7D(t)I_i(t)%7D%7BN_i(1%2B%5Cbf%7B%7BK_a%7D%5E%7BI_F%7D(p)%7D%5Ccdot%5Cbf%7B%7BM_a%7D%5E%7BI_F%7D%7D)%7D%5Cright)#0>

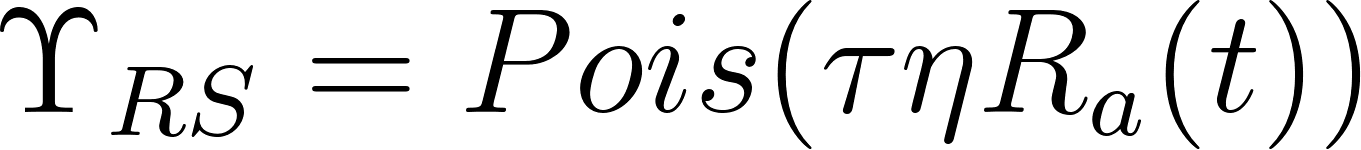
<https://www.codecogs.com/eqnedit.php?latex=%5CUpsilon_%7BRS%7D%20%3D%20Pois(%5Ctau%5Ceta%20R_a(t))#0>

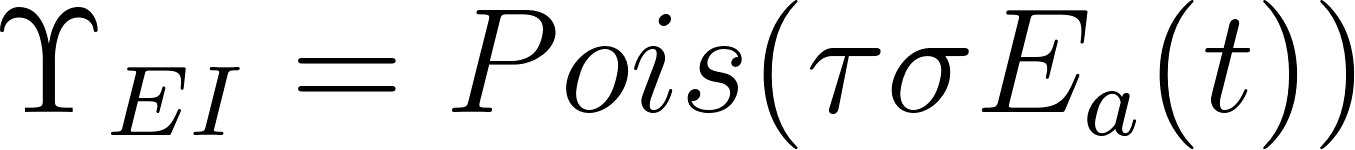
<https://www.codecogs.com/eqnedit.php?latex=%5CUpsilon_%7BEI%7D%20%3D%20Pois(%5Ctau%20%5Csigma%20E_a(t))#0>

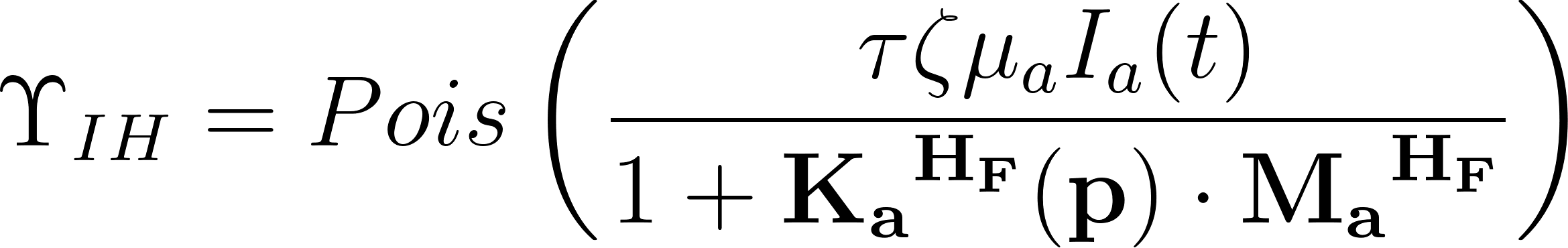
<https://www.codecogs.com/eqnedit.php?latex=%5CUpsilon_%7BIH%7D%20%3D%20Pois%5Cleft(%5Cfrac%7B%5Ctau%20%5Czeta%5Cmu_a%20I_a(t)%7D%7B1%2B%5Cbf%7B%7BK_a%7D%5E%7BH_F%7D(p)%7D%5Ccdot%5Cbf%7B%7BM_a%7D%5E%7BH_F%7D%7D%7D%5Cright)#0>

For each scenario projection, we run 1,000 stochastic simulations. We compute the seven-day rolling averages of projected hospitalizations and deaths and summarize their evolving distributions using the 0.025, 0.50, and 0.975 quantiles for each day (parameters are included in **Table S.B.3**).

#### **B7. Model parameters and fitting process**

We iteratively calibrate the transmission rate  and the age-specific infections to hospitalization rate  using least squares fitting. The fitting period spans from October 1, 2022, to April 29 2023. We initially set the  equal to published value (9), and then repeatedly (i) estimate  by fitting the models to incident hospitalization time series data(10), (ii) estimate the  by fitting the models to age-specific cumulative hospitalizations(11, 12). We repeat this two-step fitting process until the projected distribution of incident hospitalization across age groups is within 2% of the observed distribution. The transmission rate () is subject to the effects of seasonality, which fluctuates over time and is factored into  giving the time dependent transmission rate:

The seasonality magnitude (**Support Information B.4**) in the specific humidity parameter,  is estimated simultaneously along with the transmission rate. The rest of the parameters are included in the **Table S.B.3.**

**Table S.B.3.** Influenza model parameters.

| **Parameters** | **Value** | **Source** |
| --- | --- | --- |
| : US population | 334,994,511 | Ref. (13) |
| : infectious recovery rate | 0.25 | Ref. (14) |
| : hospitalized recovery rate | 0.17 | Ref. (15) |
| : transition rate out exposed | 0.5 | Ref. (16) |
| : infection hospitalization rate (IHR), age specific (2017-18 season) | [0.0378, 0.0091, 0.0025, 0.0038, 0.0105, 0.0680]* | Initialized from Ref. (9), then calibrated by fitting to Ref. (12) |
| : symptomatic to hospitalization transition rate | 0.25 | Ref. (17) |
| : in-hospital mortality rate, age specific | [0.0347, 0.0106, 0.0106, 0.0144, 0.0347, 0.0843]* | Least squares fitting to Ref. (18) assuming proportional to Ref. (19) |
| : hospitalization to death transition rate | 0.17 | Ref. (20) |
| : waning rate of immunity against infections following infection | ln(2)/(18*30) (18 months half-life) | Ref. (17, 21, 22) |
| : waning rate of immunity against infections following vaccine | ln(2)/(6*30) (6 months half-life) | Ref. (17, 21, 23) |
| : waning rate of immunity against hospitalizations following infection | ln(2)/(18*30) (18 months half-life) | Ref. (17, 21, 22) |
| : waning rate of immunity against hospitalizations following vaccine | ln(2)/(6*30) (6 months half-life) | Ref. (17, 21, 23) |
| : reduction in infection risk due to vaccine, age specific | [43%, 43%,43%, 21%, 21%, 0%]* | Ref. (2, 24) |
| : reduction in hospitalization risk due to vaccine, age specific | [68%, 68%, 68%, 23%, 23%, 41%]* | Ref. (3) |
| : transmission rate | *N*( = 0.0493,  = 0.0112) | Least squares fitting to Ref. (10) |
| : seasonality magnitude | 0.0119 | Least squares fitting to Ref. (6) |

***Age groups: [0-4, 5-11, 12-8, 19-49, 50-64, 65+]

**Table S.B.4.** For each parameter, the table provides the final calibrated value as well as the starting value and search range (in square brackets) used for calibration.

| **Parameters** | **Calibrated value** | **Starting value and range** |
| --- | --- | --- |
| : transmission rate | 0.0493 | 0.05 [0,1] |
| : seasonality magnitude | 0.0119 | 0.5 [0,1] |
| Initial proportion of the population in the infectious compartment | 0.0007% | 0.005% [0, 0.2%] |
| Initial proportion of the population in the exposed compartment | 0.0075% | 0.005% [0, 0.2%] |
| Initial proportion of the population in the hospitalized compartment | 0.0037% | 0.005% [0, 0.2%] |
| Initial percentages of population with immunity against infection | 33% | 25% [0, 50%] |
| Initial percentages of population with immunity against hospitalization | 33% | 25% [0, 50%] |
| Age-specific proportions of the initial infectious, exposed, and hospitalized compartments | [1%,1%,1%, 1%, 1%, 95%]* | 16.7% [0%-100%]  for all six age groups |
| Age-specific proportions of the initial population-level immunity | [18%, 6%, 6%, 8%, 18%, 44%] | 16.7% [0%-100%]  for all six age groups |

***Age groups: [0-4, 5-11, 12-18, 19-49, 50-64, 65+]

### **Supplementary Material C: Data**

#### **C1. 2022-2023 influenza vaccination coverage data**

In the 2022-2023 season,173.37 million doses of the influenza vaccine were distributed for the 2022-2023 US influenza season (25), which covers 49.3% of the population older than 6 months in the US (26) (**Table S.C.1**). Among children aged 6 months through 17 years, coverage was 57.8%, and among adults aged 18 years and older, it was 46.9%.

**Table S.C.1: 2022-2023 influenza vaccination coverage and hypothetical scenario if 70% of the US population had been vaccinated.**

|  | **Age group** | | | | |
| --- | --- | --- | --- | --- | --- |
|  | **0-17** | **18-49** | **50-64** | **65+** | **Total** |
| **2022-2023 Vaccine administered** | 37,441,604 | 55,805,434 | 35,589,356 | 44,533,894 | 173,370,290 |
| **2022-2023 Vaccine Coverage** | 57.8% | 35.2% | 50.1% | 69.7% | 49.3% |
| **Additional doses administered** | 9,276,975 | 53,311,336 | 13,938,575 | 1,041,728 | 77,568,614 |
| **Increase in vaccine coverage** | 12.2% | 34.8% | 19.9% | 0.3% | 20.7% |

There is some heterogeneity in vaccination coverage at the state level. The New England area has significantly higher vaccination coverage than other areas, with rates such as 65.42% in Rhode Island, 64.05% in Massachusetts, and 60.43% in Connecticut. In contrast, the Southeastern area has significantly lower vaccination coverage, with rates such as 37.65% in Florida, 42.73% in Georgia, and 42.66% in Louisiana.

**Figure S.C.1**: **Vaccination coverage percentages in different states in the US, until April 29, 2023.**

#### **C2. 2022-2023 influenza season hospital admission data**

From October 1, 2022, to April 29, 2023, there were a total of 209,972 influenza hospital admissions reported by the US Department of Health and Human Services(27). There is some heterogeneity in hospital admissions at the state level. Some southern states, such as Oklahoma (166.30 per 100,000), New Mexico (132.23 per 100,000), and Arkansas (117.09 per 100,000), experienced higher rates of influenza hospital admissions than other states. In contrast, Utah had the lowest influenza hospital admission rate in the 2022-2023 season, with only 18.17 per 100,000.

**Figure S.C.2: The overall number of hospital admissions per 100 thousand population for the 2022-2023 Influenza season in the different states in the US, spanning from October 1, 2022, to April 29, 2023.**

#### **C3. Absolute humidity data**

We incorporated absolute humidity-driven seasonality as a seasonal forcing parameter in our influenza model. We collected absolute humidity data from 2006 to 2020 from the National Oceanic and Atmospheric Administration (NOAA). Then, we calculated the average value for each day over these 15 years (**Figure S.C.3**) and assumed it to be the same value for the 2022-2023 season(6).

**Figure S.C.3. Absolute humidity data of the United States stratified to the day of a year.** Daily average of the absolute humidity data from 2006 to 2020.

**Reference**

1.	Website. Available at: [CDC. Flu vaccine provided substantial protection this season. Centers for Disease Control and Prevention https://www.cdc.gov/flu/spotlights/2022-2023/flu-vaccine-protection.htm (2023).](about:blank)

2.	E. A. Belongia, *et al.*, Variable influenza vaccine effectiveness by subtype: a systematic review and meta-analysis of test-negative design studies. *Lancet Infect. Dis.* **16**, 942–951 (2016).

3.	CDC, Flu vaccine provided substantial protection this season. *Centers for Disease Control and Prevention* (2023). Available at: <https://www.cdc.gov/flu/spotlights/2022-2023/flu-vaccine-protection.htm> [[Accessed 4 October 2023].](http://paperpile.com/b/0zr3AJ/7NpJu)

4.	Home - flu scenario modeling hub. Available at: <https://staging-extended.fluscenariomodelinghub.org/viz.html> [[Accessed 9 May 2024].](http://paperpile.com/b/0zr3AJ/Mysc)

5.	K. Bloom-Feshbach, *et al.*, Latitudinal variations in seasonal activity of influenza and respiratory syncytial virus (RSV): a global comparative review. *PLoS One* **8**, e54445 (2013).

6.	R. D. Cotton, National Oceanic and Atmospheric Administration and the environment. *Arch. Environ. Health* **22**, 404–405 (1971).

7.	[K. Prem, A. R. Cook, M. Jit, Projecting social contact matrices in 152 countries using contact surveys and demographic data. *PLOS Computational Biology* [Preprint] (2017). Available at:](http://paperpile.com/b/0zr3AJ/2c9g7) <http://dx.doi.org/10.1371/journal.pcbi.1005697>.

8.	S. J. Fox, *et al.*, Real-time pandemic surveillance using hospital admissions and mobility data. *Proc. Natl. Acad. Sci. U. S. A.* **119** (2022).

9.	M.-J. Yang, *et al.*, Influenza Vaccination and Hospitalizations Among COVID-19 Infected Adults. *J. Am. Board Fam. Med.* **34**, S179–S182 (2021).

10.	U.S. Department of Health, Human Services, COVID-19 Reported Patient Impact and Hospital Capacity by State Timeseries (RAW). Deposited 14 December 2020.

11.	N. J. Marshall, *et al.*, Influence of digital intervention messaging on influenza vaccination rates among adults with cardiovascular disease in the United States: Decentralized randomized controlled trial. *J. Med. Internet Res.* **24**, e38710 (2022).

12.	R. Garten, *et al.*, Update: Influenza Activity in the United States During the 2017-18 Season and Composition of the 2018-19 Influenza Vaccine. *MMWR Morb. Mortal. Wkly. Rep.* **67**, 634–642 (2018).

13.	U.S. Census Bureau, Explore census data. Available at: <https://data.census.gov/> [[Accessed 13 October 2023].](http://paperpile.com/b/0zr3AJ/ovkwM)

14.	S. Ghebrehewet, P. MacPherson, A. Ho, Influenza. *BMJ* **355**, i6258 (2016).

15.	Inpatient Hospital Stays and Emergency Department Visits Involving Influenza, 2006-2016 #253. Available at: <https://hcup-us.ahrq.gov/reports/statbriefs/sb253-Influenza-Hospitalizations-ED-Visits-2006-2016.jsp> [[Accessed 4 October 2023].](http://paperpile.com/b/0zr3AJ/t8WQB)

16.	Influenza (seasonal). Available at: <https://www.who.int/news-room/fact-sheets/detail/influenza-(seasonal)> [[Accessed 4 October 2023].](http://paperpile.com/b/0zr3AJ/UnSmf)

17.	M. J. Memoli, *et al.*, Influenza A Reinfection in Sequential Human Challenge: Implications for Protective Immunity and “Universal” Vaccine Development. *Clin. Infect. Dis.* **70**, 748–753 (2020).

18.	National center for health statistics mortality surveillance system. Available at: <https://gis.cdc.gov/grasp/fluview/mortality.html> [[Accessed 2 August 2023].](http://paperpile.com/b/0zr3AJ/aerW2)

19.	I. Giacchetta, C. Primieri, R. Cavalieri, A. Domnich, C. de Waure, The burden of seasonal influenza in Italy: A systematic review of influenza-related complications, hospitalizations, and mortality. *Influenza Other Respi. Viruses* **16**, 351–365 (2022).

20.	Y. Xie, T. Choi, Z. Al-Aly, Risk of Death in Patients Hospitalized for COVID-19 vs Seasonal Influenza in Fall-Winter 2022-2023. *JAMA* **329**, 1697–1699 (2023).

21.	J. M. Ferdinands, *et al.*, Intraseason waning of influenza vaccine protection: Evidence from the US Influenza Vaccine Effectiveness Network, 2011-12 through 2014-15. *Clin. Infect. Dis.* **64**, 544–550 (2017).

22.	R. G. Woolthuis, J. Wallinga, M. van Boven, Variation in loss of immunity shapes influenza epidemics and the impact of vaccination. *BMC Infect. Dis.* **17**, 632 (2017).

23.	A. Domnich, *et al.*, Waning intra-season vaccine effectiveness against influenza A(H3N2) underlines the need for more durable protection. *Expert Rev. Vaccines* **23**, 380–388 (2024).

24.	M. T. Osterholm, N. S. Kelley, A. Sommer, E. A. Belongia, Efficacy and effectiveness of influenza vaccines: a systematic review and meta-analysis. *Lancet Infect. Dis.* **12**, 36–44 (2012).

25.	Influenza Vaccine Doses Distributed, United States. (2024). Available at: <https://www.cdc.gov/flu/fluvaxview/dashboard/vaccination-doses-distributed.html> [[Accessed 5 March 2024].](http://paperpile.com/b/0zr3AJ/k0990)

26.	Flu vaccination coverage, United States, 2022–23 influenza season. (2023). Available at: <https://www.cdc.gov/flu/fluvaxview/coverage-2223estimates.htm> [[Accessed 17 April 2024].](http://paperpile.com/b/0zr3AJ/7MBU)

27.	U.S. Department of Health, Human Services, COVID-19 reported patient impact and hospital capacity by state timeseries (RAW). Deposited 14 December 2020.
